# Metabolomic Responses to External and Internal Environmental Exposures: Evidence of Lipid and Energy Metabolism Disruption in the Estonian Biobank

**DOI:** 10.64898/2026.03.17.26347937

**Authors:** Max J. Oosterwegel, Roel Vermeulen, Estonian Biobank research team, Kees de Hoogh, Liis Hiie, Tõnu Esko, Jelle Vlaanderen, Jaanika Kronberg

## Abstract

**Objective:** To investigate associations between long-term environmental exposures, both external (ambient air pollution and built environment) and internal (circulating anthropogenic chemicals), and the human plasma metabolome, with the aim of generating biologically plausible hypotheses about affected metabolic pathways.

**Methods:** We analyzed plasma from 989 Estonian Biobank participants using untargeted LC-HRMS (Metabolon HD4). External exposures (*PM*_2.5_, *PM*_10_, *NO*_2_, ozone and built-environment metrics) were assigned using spatiotemporally resolved models developed in the EXPANSE project. Internal exposures were defined as ubiquitous anthropogenic compounds detected in the same metabolomics dataset. Associations between exposures and individual metabolites were quantified using left-censored regression models and then mapped to metabolite classes (Metabolon) and KEGG pathways. For enrichment analyses, one-sided Kolmogorov-Smirnov tests were applied to external exposures and Fisher’s exact tests to internal exposures. False discovery rate was controlled at 1% per exposure and database.

**Results:** External air pollutants exhibited distinct metabolic patterns: Higher *NO*_2_ exposure was associated with enrichment of metabolites involved in tyrosine metabolism; higher ozone with monohydroxy and dicarboxylate fatty acids (consistent with lipid peroxidation); and higher *PM*_2.5_ with acyl-carnitine subclasses and carbohydrate metabolism (glycolysis / gluconeogenesis / pyruvate). Built-environment associations were heterogeneous across metabolites and pathways. Internal anthropogenic chemicals showed broader metabolic associations than external exposures, involving a larger number of metabolites and metabolic classes. PFAS (PFOA, PFOS) were associated with long-chain polyunsaturated fatty acids (n3/n6) and lysophospholipids. Associations with 4-hydroxychlorothalonil, a fungicide, pointed to androgenic steroid metabolites and alpha-linolenic acid metabolism. The phenolic 2,4-di-tert-butylphenol, a plastic associated chemical, showed widespread associations with lipid classes, suggesting disruption of membrane remodeling and fatty acid handling.

**Conclusion:** Long-term environmental exposures, both external and internal, are measurably reflected in the human plasma metabolome. Across exposure domains, recurrent signals involved lipid metabolism, membrane composition, and oxidative stress–related pathways, highlighting these as common biological targets of environmental exposures. The findings generate testable hypotheses, including nitrosative stress–related alterations for *NO*_2_, lipid peroxidation for ozone, energy-metabolism perturbations for *PM*_2.5_, potential endocrine activity for chlorothalonil metabolites, and possible obesogenic effects of 2,4-di-tert-butylphenol.

## 1 INTRODUCTION

Genetic factors explain only a minority of chronic disease burden, underscoring the central role of environmental exposures in shaping human health [Miller and Banbury Exposomics Consortium, 2025, Vermeulen et al., 2020]. Understanding how diverse exposures, i.e. the exposome, interact with human biology at a molecular level is essential for elucidating disease mechanisms and eventually informing public health strategies.

Exposomics has emerged as a discovery-driven framework for systematically studying these influences using high-dimensional molecular data [Safarlou et al., 2024]. For example, transcriptomic studies have identified gene-expression signatures associated with long-term fine particulate matter (*PM*_2.5_) exposure [Vlaanderen et al., 2022], while epigenomic analyses have revealed differential DNA methylation patterns related to pesticide use [van der Plaat et al., 2018].

Among omic approaches, the untargeted liquid chromatography-high-resolution mass spectrometry (LC-HRMS) metabolomics is particularly useful for exposomics because it can simultaneously capture a comprehensive snapshot of both exposure biomarkers and internal biological responses. High-resolution metabolomics has successfully detected specific exposure-related compounds, such as known benzene metabolites in occupationally exposed workers, while also revealing downstream perturbations in mitochondrial and energy metabolism that mirror observations from animal models [Rothman et al., 2021]. Other studies have linked known and unknown metabolites of trichloroethylene to pathways suggestive of autoimmunity and renal toxicity [Walker et al., 2016], and a growing body of evidence connects per- and polyfluoroalkyl substances (PFAS) to disruptions in lipid metabolism and metabolic stress [India-Aldana et al., 2023]. Together, these findings highlight that LC-HRMS metabolomics can bridge external exposures with internal biological processes, making it a powerful tool for exposome-wide investigations.

Despite these advances, most metabolomics studies have been relatively small, limited to short-term exposures, or focused on a narrow range of environmental factors [Ginos et al., 2024]. Moreover, the exposome encompasses both external exposures (e.g. pollutants, features of built and social environments) and internal exposures (e.g. circulating anthropogenic chemicals), yet these domains are rarely examined together. External exposures may directly perturb endogenous metabolism, but they may also correlate with internal chemical burdens through shared sources, behaviors, or environmental contexts. The degree to which these two components of the exposome jointly shape the human metabolome remains largely unexplored.

To address this gap, we leverage data from the Estonian Biobank to examine how both external exposures (air pollution and built-environment characteristics) and internal exposures (circulating anthropogenic chemicals) relate to the plasma metabolome. We characterize external exposures using harmonized, spatiotemporally resolved air pollution and built-environment surfaces developed within the EXPANSE consortium [Vlaanderen et al., 2021, de Hoogh et al., 2025], and we derive internal exposures directly from the same LC-HRMS dataset by identifying circulating ubiquitous anthropogenic chemicals with high analytical confidence. By jointly examining these external and internal components of the exposome in relation to the plasma metabolome, we aim to generate biologically plausible hypotheses about affected metabolic pathways and their potential relevance for environmental health.

## 2 MATERIALS AND METHODS

Figure 1 shows the general workflow of analyses.

**Figure 1.**
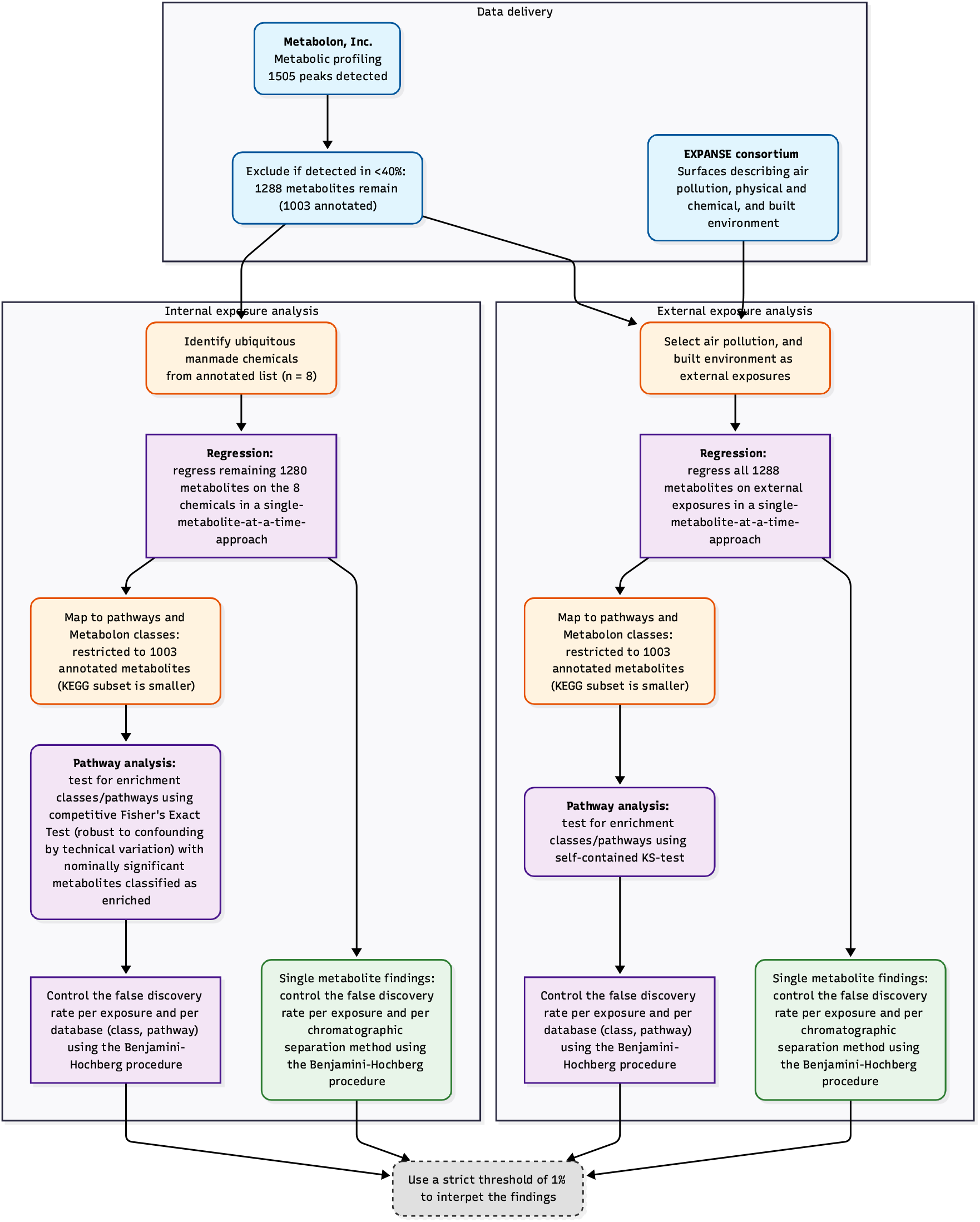
Analysis workflow

### 2.1 Study population

The Estonian Biobank has been described in detail before [Milani et al., 2025, Leitsalu et al., 2015]. In brief, the biobank is a volunteer-based adult cohort of over 212,000 participants of approximately 20% of the Estonian adult population. Estonian Biobank recruitment was extended over a period of 25 years, however over 90% of subjects were recruited in two big waves: 50 000 subjects in 2003-2006 and 150 000 in 2018-2019. The metabolomics dataset of 991 samples has been described in a previous publication [Yu et al., 2025a]. In brief, metabolomic analysis was performed in samples from 723 individuals with predicted loss of function variants in genes associated with diseases, and 268 random controls. Two control samples were excluded due to insufficient plasma volume.

### 2.2 Metabolomic analyses

EDTA plasma samples were sent to Metabolon, Inc. (Morrisville, NC, USA) and analyzed using their HD4 LC-HRMS platform. In brief, this platform leverages four distinct chromatographic separations: C18 reversed-phase method with negative ion mode, a HILIC method for polar compounds, and two C18 reversed-phase methods with positive ion mode, one optimized for more hydrophilic species and another for more hydrophobic species. Metabolomic features from the platform were identified by matching them to Metabolon’s proprietary (reference) library.

### 2.3 Metabolic databases

For pathway enrichment analyses, we linked the identifiers (PubChem [Kim et al., 2025], ChemSpider [Williams et al., 2010], KEGG [Kanehisa and Goto, 2000], HMDB [Wishart et al., 2007] – provided by Metabolon) of the metabolic data to the public KEGG metabolite pathway databases using RaMP-DB 2.015 [Braisted et al., 2023]. Pathways were included in the enrichment analysis only if at least five detected metabolites mapped to them. The subclass description (Table S4) is a label assigned by Metabolon that groups metabolites based on shared structural or biochemical characteristics (e.g., bile acids, fatty acids, amino acid derivatives). Unlike KEGG pathway annotations, which were available only for a subset of metabolites, these subclass descriptions were available for all annotated metabolites and assigned a single class to each feature. Tables S4 and S9 describe all classes and pathways included in the analysis.

### 2.4 Exposures

Environmental exposure surfaces were created by the EXPANSE consortium and have been described before [de Hoogh et al., 2025, Shen et al., 2024]. In summary, the consortium developed harmonized, Europe-wide spatially and temporally varying layers for major air pollutants (*PM*_2.5_, *PM*_10_, ozone, *NO*_2_), physical-chemical variables (daily temperature, annual-average road-traffic noise) and built environment indicators (green, blue, and grey spaces (imperviousness), walkability, light-at-night, and urbanicity).

In this analysis, we used these layers to estimate the average air pollution exposure over the 12 months preceding the blood draw (including the month of blood collection itself) based on the participant’s residential self-reported locations at recruitment. For the built environment domain, we linked the surface to the most recent year before the blood collection with data available. For more stable surfaces such as blue space and urban/rural indicators the closest surface in time irrespective of the temporal ordering was used. We excluded imperviousness (due to high missingness), distance to green space, and light at night from the analysis of this Estonian dataset. Distance to green space was excluded because the Urban Atlas variable only covered cities, excluding rural areas, and the Corine-based variable missed important accessible green areas such as university grounds. We found that NDVI and MSAVI better captured actual greenness and provided complete country coverage. For variables (MSAVI, NDVI) with multiple buffer sizes, we selected a 500m buffer a priori to align with existing literature (e.g. Yu et al. (2025) [Yu et al., 2025b]).

To characterize internal exposures, we screened the list of annotated Metabolon features for ubiquitous anthropogenic organic chemicals of concern. Only compounds detected with the highest level of analytical confidence and with *<* 10% nondetects were retained. Details on preprocessing these biomarkers are described in S1 Supplementary Methods I.

### 2.5 Statistical methods

#### 2.5.1 Principal components of external exposures

To account for the correlated structure of the air pollution variables (Figure S1) and the built-environment variables (Figure S2), we applied principal component analysis (PCA) separately within each domain [de Bont et al., 2023]. For air pollution, the input variables were *PM*_2.5_, *PM*_10_, ozone and *NO*_2_; for built-environment features, the inputs were distance to the nearest inland freshwater, distance to the nearest sea/ocean, distance to the nearest blue space, modified soil-adjusted vegetation index (MSAVI), and normalized difference vegetation index (NDVI). Within each domain, we retained the number of components required to explain a combined 80% of the total variance (N=2 for air pollution, and N=3 for built environment).

#### 2.5.2 Associations of exposures with single metabolites

Metabolites detected in *<* 40% of the samples were excluded from analysis. We then regressed each metabolite on each exposure in a one-metabolite/one-exposure-at-a-time framework. To handle missingness the intensity of a metabolite was modelled as a potentially left-censored (at the minimum detected value of a batch) natural log-transformed variable:

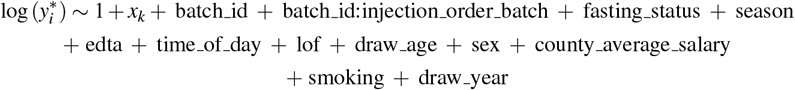

For all metabolites *i*, and all exposures *k*.

Smoking was coded as (never, current, former), county average salary was defined using data from Statistics Estonia and categorized as high, low, or intermediate based on the participant’s county of residence, sex (male, female), age at sample collection (rounded to years), sample draw year as continuous variable, loss-of-function (LOF) as a binary indicator of sample selection group, fasting status indicates that fasting was required by protocol and coded as a binary indicator, season (winter (December – February), spring (March – May)), summer (June – August), autumn (September – November)), and time of day (before 10, between 10 and 15, after 15 o’clock). EDTA intensity levels (as measured by the LC-MS platform) were added as an adjustment variable because slight variations in EDTA intensities were observed across samples (see ‘Supplementary Methods I’ on how the EDTA variable was preprocessed).

#### 2.5.3 FDR adjustment for single associations

We controlled the false discovery rate (FDR) using the Benjamini-Hochberg (BH) procedure at a conservative 1% per exposure (*PM*_2.5_, ozone, PFOA etc.), and per chromatographic separation method to prioritize the identification of highly robust associations and minimize the potential for type I errors (false positives). We implemented chromatographic method specific FDR adjustment due to anticipated differences in signal-to-noise ratios arising from potential differences in technical variation and the distinct chemical space covered by each method.

#### 2.5.4 Pathway enrichment analysis: single metabolites (external exposures)

To identify which biological pathways were potentially associated with exposures from the air pollution and built environment domain, we employed a one-sample one-sided Kolmogorov-Smirnov (KS) test with a uniform reference distribution to the results of the single metabolite at a time approach. This self-contained test evaluates pathway enrichment by assessing whether the p-values for metabolites within a pathway show an excess of small values relative to the null expectation of no association between the exposure and the pathway [de Leeuw et al., 2016].

#### 2.5.5 Pathway enrichment analysis: single metabolites (internal exposures)

Due to concern of confounding by technical variation (details in S1 Supplementary Methods I), we ran a competitive test to test which pathways are associated with internal exposures using Fisher’s exact test. A competitive test tests whether the proportion of statistically significant hits is greater inside a pathway than outside (‘background’) and thus adjusts for a potential inflation of hits due to confounding by technical variation. Nominally statistically significant metabolites (*p <* 0.05) were considered hits in this analysis.

#### 2.5.6 FDR pathways

We controlled the FDR (using the BH procedure) at a conservative 1% per exposure (*PM*_2.5_, ozone, PFOA etc.), per database (Metabolon classes, KEGG) to prioritize the identification of highly robust associations and minimize the potential for type I errors (false positives).

## 3 RESULTS

### 3.1 Study population

989 participants were included. Detailed information on the subjects can be found in (Table 1). In Brief, samples were collected between 2002 and 2019. The average age was 45 (youngest 18, oldest 93). 63% of the participants were female. 47% lived in cities while 33% lived in rural areas. The average air pollution levels per year for the home address for participants included in our dataset is displayed in Figure S3. Median concentrations were 7.8 µg*/*m^3^ for *PM*_2.5_, 9.7 µg*/*m^3^ for *NO*_2_, 15 µg*/*m^3^ for *PM*_10_ and 64.3 µg*/*m^3^ for ozone. *PM*_2.5_ and *NO*_2_ levels fell below new EU guidelines and approached WHO recommendations (*PM*_2.5_: 5 µg*/*m^3^; *PM*_10_: 15 µg*/*m^3^; *NO*_2_: 10 µg*/*m^3^; ozone: 60 µg*/*m^3^ peak season). Across all categories, pollutant concentrations decreased over the course of the study. No discernible temporal trends in vegetation indices and other built environment variables were observed.

**Table 1.** Participants included in the study. LOF = selected for loss of function individual in previous study; MSAVI = modified soil adjusted vegetation index; NDVI = Normalized Difference Vegetation Index; BSS = distance to nearest sea/ocean; BSI = distance to nearest inland freshwater; BSW = distance to nearest blue space; UR = Population density as focal mean within a 1500m square.

| (a) Covariates |  | (b) External exposures |  |
| --- | --- | --- | --- |
| N | 989 | *NO*<sub>2</sub> [µg/m<sup>3</sup>] | 9.7 (5.7, 14.6) |
| Age | 45 (32, 59) | Unknown | 16 |
| Sex |  | OZO [µg/m<sup>3</sup>] | 64.3 (61.1, 66.7) |
| Female | 624 (63.1%) | Unknown | 16 |
| Male | 365 (36.9%) | *PM*<sub>10</sub> [µg/m<sup>3</sup>] | 15.01 (13.46, 16.87) |
| Smoking |  | Unknown | 16 |
| Current | 182 (18.4%) | *PM*<sub>2.5</sub> [µg/m<sup>3</sup>] | 7.83 (6.70, 9.30) |
| Former | 209 (21.1%) | Unknown | 81 |
| Never | 492 (49.7%) | MSAVI | 7,436 (6,786, 7,958) |
| Unknown | 106 (10.7%) | Unknown | 18 |
| BMI | 26.1 (22.5, 30.0) | NDVI | 5,922 (5,147, 6,617) |
| Unknown | 106 (10.7%) | Unknown | 18 |
| Education |  | BSI [m] | 1,706 (707, 3,400) |
| Low | 75 (7.6%) | Unknown | 15 |
| Intermediate | 448 (45.3%) | BSS [m] | 34,800 (2,800, 107,579) |
| High | 359 (36.3%) | Unknown | 15 |
| Unknown | 107 (10.8%) | BSW [m] | 1,290 (608, 2,720) |
| Year of blood draw | 2011 (2009, 2018) | Unknown | 15 |
| Time of day |  | UR | 10 (1, 30) |
| After 15 o'clock | 191 (19.3%) | Unknown | 15 |
| Before 10 o'clock | 217 (21.9%) |  |  |
| Between 10 and 15 o'clock | 475 (48.0%) |  |  |
| Unknown | 106 (10.7%) |  |  |
| Region |  |  |  |
| City | 412 (41.7%) |  |  |
| Rural area | 293 (29.6%) |  |  |
| Town | 94 (9.5%) |  |  |
| Unknown | 190 (19.2%) |  |  |
| Fasted | 694 (70.2%) |  |  |
| LOF |  |  |  |
| 0 | 266 (26.9%) |  |  |
| 1 | 723 (73.1%) |  |  |

<sup>1</sup>(%); Median (Q1, Q3)

### 3.2 PCA air pollution and built environment

Detailed loadings for all principal components are provided in S2 Supplementary Information I. In summary, two principal components of the air pollution domain explained *>* 80% of the variation among the air pollutants. The first component explained 62% of the variation and corresponded to time and places with higher ozone levels, but lower levels of *PM*_10_, *PM*_2.5_ and *NO*_2_. The second principal component explained 29% of the variation and corresponded to time and places with higher *NO*_2_ levels, but lower *PM*_10_, *PM*_2.5_ and ozone levels.

Three principal components of built environment explained *>* 80% of the variation among built environment. The first component corresponded to rural and natural environments (49%), while the second seemed to describe areas far from the coast but not necessarily close to inland lakes or rivers (28%), and the third component (14%) was dominated by a high positive loading for distance to the sea/ocean (BSS). Given that Tallinn, Estonia’s capital city and major economic hub, is located on the coast, this component may not only represent coastal proximity but could also serve as a proxy for the unique socio-environmental characteristics associated with the capital region.

### 3.3 Assessment of blood metabolome

In total, 1505 compounds were detected by the LC-HRMS platform. After excluding compounds with insufficient detection rates, 1288 compounds remained for analysis (see S3 Supplementary Information II for a breakdown by column and ionization mode). These 1288 compounds comprised of 70 Metabolon classes and 41 KEGG pathways that met the minimum membership threshold.

#### 3.3.1 Assessment of internal exposures

Eight ubiquitous anthropogenic chemicals were identified: the phenolic compound 2,4-di-tert-butylphenol (a suspected endocrine disruptor), three PFAS compounds (PFOA, PFOS, and PFHxS), 2-naphthol sulfate (a suspected carcinogen), 4-hydroxychlorothalonil (a metabolite of the fungicide chlorothalonil, itself an endocrine disruptor and suspected carcinogen), and two additional chlorinated compounds (3,5-dichloro-2,6-dihydroxybenzoic acid and 4-chlorobenzoic acid).

### 3.4 Associations of exposures with single metabolites

#### 3.4.1 External exposures: air pollution and built environment

The associations between external exposures and individual metabolites are shown in volcano plots in Figure S4 and Figure S5, and statistically significant metabolites are listed in Table S3. At an FDR of 1%, *NO*_2_ was associated with 12 metabolites, ozone with 15 metabolites, and *PM*_2.5_ with 13 metabolites, while *PM*_10_ showed no significant associations at this threshold. For the air-pollution principal components, PC1 was associated with five metabolites and PC2 with eight metabolites (Table S3).

Among the metabolites associated with *NO*_2_, ozone and air-pollution PC1, several were anthropogenic compounds detected in the metabolomics data, including PFHxS, PFOS, 4-hydroxychlorothalonil, 2-naphthol sulfate and 3,5-dichloro-2,6-dihydroxybenzoic acid. The remaining associated metabolites were endogenous compounds, indicating that external air pollution is linked both to internal anthropogenic chemical levels and to broader endogenous metabolic perturbations.

Built-environment exposures showed heterogeneous associations with the metabolome (Table S5). Distance to the sea/ocean exhibited the largest signal, with 41 associated metabolites. NDVI and MSAVI were each associated with the same seven metabolites, and the urban–rural classification with eight metabolites. Distance to inland freshwater was associated with two metabolites, while distance to the nearest blue space showed no significant associations. For the built-environment principal components, PC1 was associated with seven metabolites, PC2 with eight, and PC3 (interpreted as coastal proximity/capital region characteristics) with 28 metabolites (Table S3).

As with air pollution, these associations comprised both exogenous (anthropogenic) and endogenous metabolites, suggesting that features of the built environment are linked to patterns of internal exposure and metabolism. Specifically, urbanicity was associated with higher levels of 4-hydroxychlorothalonil, whereas rural areas were associated with 2-naphthol sulfate and PFHxS levels. Higher greenness indicators (MSAVI and NDVI) and higher values of PC1 showed positive associations with PFOS and PFHxS, and negative associations with 4-hydroxychlorothalonil. PC3 demonstrated negative associations with PFOS and PFHxS, while PC2 showed negative associations with 4-hydroxychlorothalonil. Regarding blue spaces, greater distance to the sea was associated with lower levels of PFOS, 4-hydroxychlorothalonil, and 3,5-dichloro-2,6-dihydroxybenzoic acid, whereas greater distance to inland water was associated with higher PFOS levels.

#### 3.4.2 Internal exposures: ubiquitous anthropogenic chemicals

The eight anthropogenic chemicals identified in the dataset showed extensive metabolic associations, substantially exceeding those observed for external environmental exposures. The phenolic compound 2,4-di-tert-butylphenol showed the broadest metabolic footprint, with 350 associated metabolites, followed by PFOA (245 metabolites), 2-naphthol sulfate (208 metabolites), PFHxS (187 metabolites), PFOS (172 metabolites), 3,5-dichloro-2,6-dihydroxybenzoic acid (136 metabolites), 4-hydroxychlorothalonil (128 metabolites), and 4-chlorobenzoic acid (85 metabolites).

These patterns indicate that the internal anthropogenic exposome is strongly embedded in the blood metabolome, with each chemical associated with wide-ranging perturbations in endogenous metabolic pathways.

### 3.5 Enrichment of exposures with Metabolon classes

#### 3.5.1 External exposures: air pollution and built environment

Significant class-level enrichments are summarized in Table 3. In brief, metabolites associated with higher *PM*_10_ and *PM*_2.5_ were enriched in long-chain saturated and monounsaturated fatty acid classes. Metabolites associated with ozone, *PM*_10_, and *PM*_2.5_ were enriched in long-chain polyunsaturated fatty acids. Metabolites linked to *PM*_10_ and *PM*_2.5_ were also enriched in the acyl choline subclass of fatty acid metabolism, whereas metabolites associated with *NO*_2_ and ozone were enriched in acyl glycine. Metabolites associated with both *NO*_2_ and ozone were enriched in the food component/plant class.

**Table 3.**
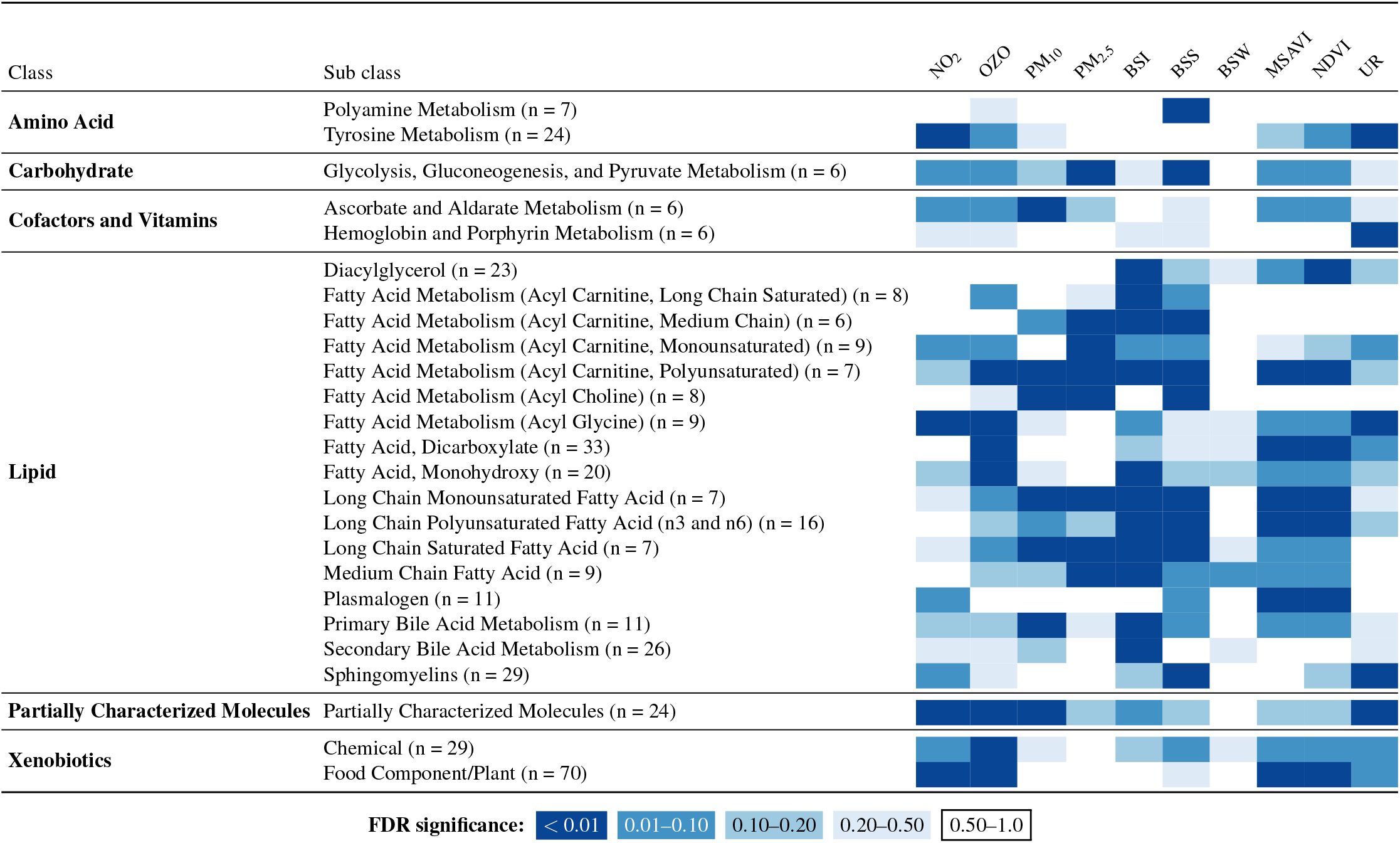
Metabolon classes significantly enriched for associations with internal exposures (25/70 classes showed significant enrichment). n = number of members pathway. MSAVI = modified soil adjusted vegetation index; NDVI = Normalized Difference Vegetation Index; BSS = distance to nearest sea/ocean; BSI = distance to nearest inland freshwater; BSW = distance to nearest blue space; UR = Population density as focal mean within a 1500m square.

Several enrichment signals were specific to individual air pollutants. Metabolites associated with *NO*_2_ were enriched for tyrosine metabolism; metabolites associated with *PM*_10_ were enriched for primary bile acid metabolism and ascorbate/aldarate metabolism; and metabolites associated with *PM*_2.5_ were enriched for acyl carnitine subclasses (monounsaturated and medium-chain) and for glycolysis, gluconeogenesis, and pyruvate metabolism. Finally, metabolites associated with ozone were enriched in the monohydroxy and dicarboxylate fatty acid classes.

For built-environment variables, metabolites associated with NDVI and MSAVI showed enrichment in several fatty acid subclasses.

#### 3.5.2 External exposures: PCs of air pollution and built environment

All metabolite classes statistically significantly enriched for a PC of external exposures are shown in Table S6. For air pollution, PC1 showed enrichment for three fatty acid metabolism subclasses (acyl carnitine, polyunsaturated; acyl choline; and acyl glycine), while PC2 showed enrichment for acyl carnitine (medium chain) and for food component/plant metabolites.

For built-environment features, several long-chain fatty acid subclasses showed enrichment for PC1 and PC3.

#### 3.5.3 Internal exposures: ubiquitous anthropogenic chemicals

Class-level enrichments for internal exposures are shown in Table 4 and the correlation structure of the anthropogenic chemicals is shown in Figure S6. Common enrichments included: 2,4-di-tert-butylphenol, 4-hydroxychlorothalonil, PFOS, PFOA compounds showed enrichment for metabolites from the long chain polyunsaturated fatty acid (n3 and n6) class. Moreover, 2,4-di-tert-butylphenol, 2-naphthol sulfate, PFOA showed enrichment for lysophospholipid metabolites, and 2,4-di-tert-butylphenol, and 4-chlorobenzoic acid showed enrichment for phosphatidylethanolamine.

**Table 4.**
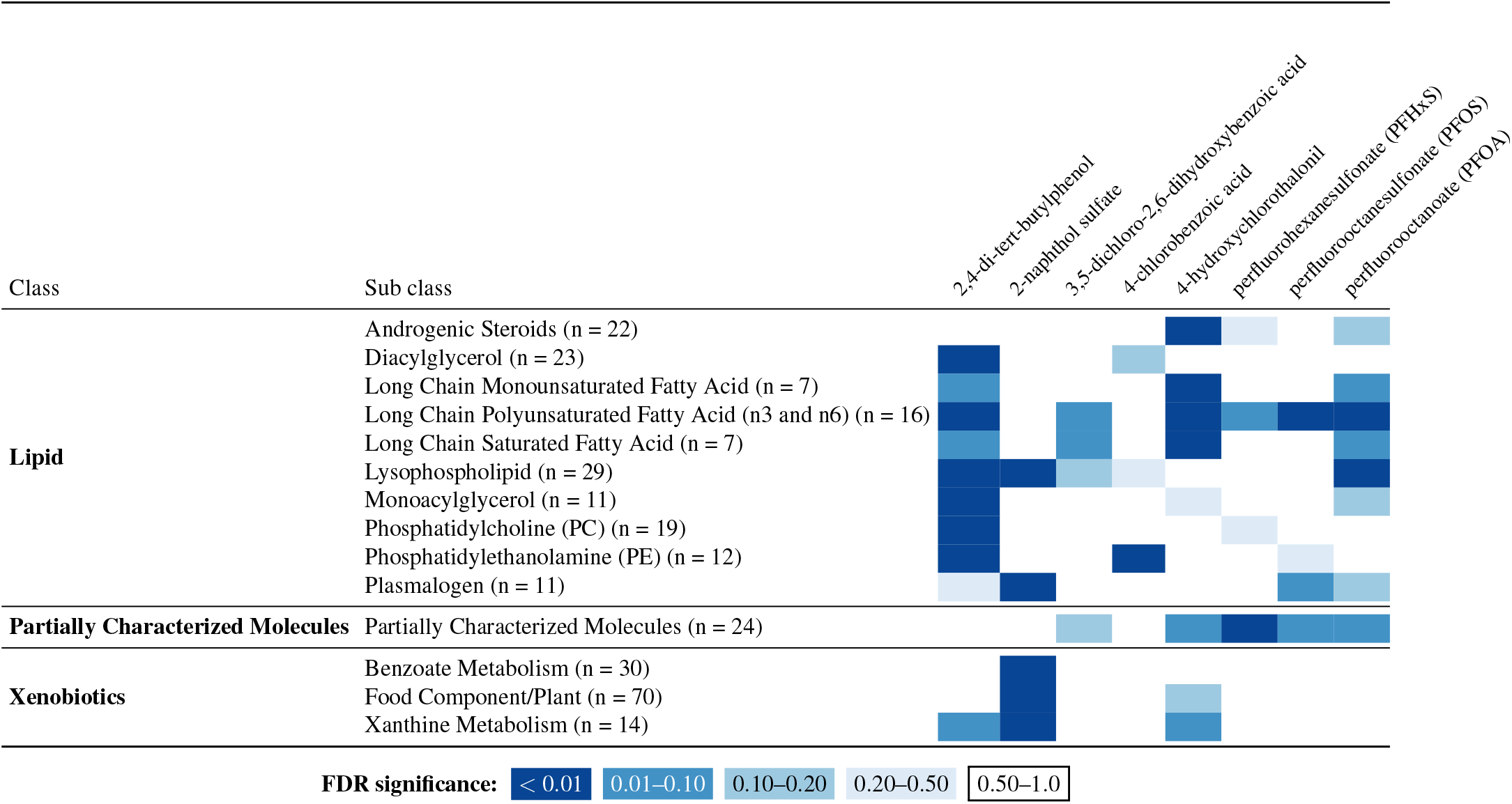
Metabolon classes significantly enriched for associations with external exposures (14/70 classes showed significant enrichment). n = number of members pathway in dataset.

There were also hits unique to single internal exposures, namely 2-naphthol sulfate showed enrichment for benzoate metabolism, plasmalogen, and xanthine metabolism; 2,4-di-tert-butylphenol to diacylglycerol, monoacylglycerol, and phosphatidylcholine metabolites, whereas 4-hydroxychlorothalonil showed enrichment for metabolites from the androgenic steroids, long chain monounsaturated fatty acid, long chain saturated fatty acids classes. 3,5-dichloro-2,6-dihydroxybenzoic acid did not cross the false discovery threshold for any metabolite class.

### 3.6 Enrichment of exposures with KEGG pathways

#### 3.6.1 External exposures: air pollution and built environment

KEGG pathway enrichment is shown in Table S8. Fewer pathways reached statistical significance compared to the Metabolon class-level results. For air pollutants, metabolites associated with *PM*_10_ showed enrichment for glycine/serine/threonine metabolism, methionine metabolism and steroid biosynthesis. Metabolites associated with *PM*_2.5_ showed a similar pattern but crossed the threshold only for starch and sucrose metabolism. Among gaseous pollutants, metabolites associated with ozone showed no enrichment for any KEGG pathways, whereas metabolites associated with *NO*_2_ showed enrichment for phosphatidylinositol phosphate metabolism.

Within the built environment domain, statistically significant enrichment was observed for distance to nearest sea/ocean (25 KEGG hits), urban–rural classification (3 hits), PC2 (13 hits) and PC3 (21 hits).

#### 3.6.2 External exposures: PCs of air pollution and built environment

No KEGG pathways were significantly enriched among metabolites associated with the air-pollution PCs. In contrast, metabolites associated with built-environment PC2 were enriched in 13 KEGG pathways, and metabolites associated with built-environment PC3 were enriched in 21 pathways.

#### 3.6.3 Internal exposures: ubiquitous anthropogenic chemicals

For internal exposures, only 4-hydroxychlorothalonil showed a significant enrichment for a KEGG pathway: alpha-linolenic acid metabolism.

## 4 DISCUSSION

In this study, we jointly examined how long-term external environmental exposures (air pollution and built environment) and internal circulating anthropogenic chemicals relate to the human plasma metabolome. By combining untargeted LC-HRMS metabolomics with high-resolution spatial exposure models, we characterized both the direct metabolic effects of internal chemicals and the broader biological responses associated with external air pollution and built-environment features. This dual-exposome approach allowed us to identify converging metabolic themes across exposure domains and generate mechanistically plausible hypotheses for future targeted studies.

Across both external and internal exposures, we consistently observed perturbations involving lipid metabolism, membrane remodeling, and oxidative stress–related pathways, suggesting these are recurrent biological targets of environmental exposures in this population.

### 4.1 External exposures: air pollution and built environment

Our findings corroborate and extend recent metabolomics evidence linking air pollution to systemic metabolic stress [Liang et al., 2023]. We replicated the association between *NO*_2_ exposure and tyrosine metabolism – a link supported by established nitrosative stress pathways where *NO*_2_ acts as a reactant in the free radical-mediated nitration of tyrosine residues [Radi, 2004]. In our study, *NO*_2_ levels were also related to the phosphatidylinositol phosphate metabolism, a pathway whose components are involved in signaling processes mediating responses to oxidative stress and inflammation thereby reinforcing the connection between traffic related oxidants and systemic metabolic stress [U.S. Environmental Protection Agency, 2020, Song et al., 2025, Chen et al., 2009]. Our observation of an association of *PM*_10_ with the methionine pathway also aligns with the review’s identification of methionine and cysteine metabolism with air pollution. However, our results did not mirror every pathway identified in this review (such as glycerophospholipid, tryptophan or pyrimidine metabolism). Such variation may be expected with differences in study time-window (mostly short-term versus long-term), study design (randomized cross-over versus observational), or environmental context, specifically the relatively low air pollution levels in Estonia.

Beyond these replicated associations, we observed specific metabolic signatures that, while less commonly reported in previous epidemiological studies, appear consistent with toxicological mechanisms of pollutant toxicity. Metabolites associated with ozone exposure were enriched in lipid peroxidation products, including monohydroxy and dicarboxylate fatty acids, reflecting the oxidative attack of this reactive pollutant on polyunsaturated fatty acids [Klaassen, 2018]. *PM*_2.5_ was associated with the broadest set of metabolic changes, with significant associations across multiple acyl carnitine classes and alterations in the glycolysis, gluconeogenesis, and pyruvate metabolism class and the starch and sucrose metabolism pathway (reflecting glucose and glycogen homeostasis) – a pattern consistent with its documented links to mitochondrial dysfunction and insulin resistance [U.S. Environmental Protection Agency, 2019]. Both *PM*_10_ and *PM*_2.5_ showed perturbations in fatty acid metabolism, though *PM*_2.5_ exhibited broader systemic effects potentially related to the deeper lung penetration and greater potential for systemic circulation [Klaassen, 2018, U.S. Environmental Protection Agency, 2019]. The significant association between *PM*_10_ and the glycine, serine, and threonine metabolism is also plausible. This pathway synthesizes glycine, a critical component of the antioxidant glutathione [Nelson and Cox, 2005], which is rapidly depleted to combat the oxidative stress induced by particulate matter [U.S. Environmental Protection Agency, 2019]. Multi-pollutant principal component analysis further supported source-related differences in metabolic signatures: contrasts between ozone-dominated versus combustion-related pollution profiles were linked to distinct fatty acid pathways, while *NO*_2_-dominated profiles were more closely related to medium-chain acyl-carnitines. Importantly, these potential metabolic perturbations were observed at air pollution levels approximating current WHO guidelines, underscoring the biological potency of even low-level exposure.

Some findings were less conventional. The association of *PM*_10_ with primary bile acid metabolism, and steroid hormone metabolism is not a classic pathway of particulate toxicity. However, given that coarse particles can be cleared via swallowing [U.S. Environmental Protection Agency, 2019], these signals – particularly bile acids – invite speculation regarding a potential ‘lung-gut-liver axis’ component to the exposure, although this remains an interpretive possibility requiring further investigation [Mazumder and Hussain, 2024, Mutlu et al., 2018]. Such a pathway provides a biologically plausible mechanism for the established epidemiological link between air pollution and metabolic disorders. For example, considering that both air pollution exposure [He et al., 2025] and bile acid are linked to the etiology of non-alcoholic fatty liver disease (NAFLD) [Yuan and Bambha, 2015], our findings may offer a ‘meet-in-the-middle’ explanation for how inhaled (and subsequently swallowed) coarse particles might drive liver pathology.

By comparison, other signals were less readily interpretable. For example, enrichment of acyl-choline metabolites for particulate matter could reflect altered membrane signaling or choline metabolism, but the mechanistic basis remains uncertain. Similarly, while *PM*_10_ showed associations with ascorbate and aldarate metabolism, the absence of a similar signal for ozone – despite its well-documented depletion of airway antioxidants [Klaassen, 2018] – shows that some expected pathways have fallen just below our stringent significance threshold, highlighting the trade-off between minimizing false positives and capturing the full biological response to air pollutant exposure.

Most built-environment associations were driven by distance to the sea and built-environment PC3, which largely captured the socio-environmental profile of the coastal/capital region (Tallinn). These signals likely reflect broader socio-environmental differences associated with the coastal/capital region rather than a direct biological effect of coastal proximity itself. (e.g., differences in lifestyle, socioeconomic conditions, urban design, occupation, diet, or co-exposures). Thus, these metabolic signatures should be interpreted as reflecting contextual environmental differences rather than causal effects of geography alone.

Similarly, associations with vegetation indices (NDVI, MSAVI) were modest and should be interpreted cautiously. In Estonia, greener areas generally coincide with lower air pollution, making it difficult to disentangle greenness from co-varying pollutant exposures. Our findings therefore do not support direct biological effects of greenness but rather suggest that vegetation may act as a proxy for broader environmental conditions.

### 4.2 Internal exposures: PFAS, chlorinated compounds, and other chemicals PFAS

We observed robust associations of PFOA and PFOS with long-chain polyunsaturated fatty acids (n-3 and n-6) and lysophospholipids, consistent with prior metabolomics studies. These findings are consistent with the literature demonstrating that PFAS disrupt fatty acid metabolism, largely through the activation of nuclear receptors like *PPAR* – *α* [Evans et al., 2022]. Furthermore, we observed a specific association between PFOA and the lysophospholipid class. While this supports the general picture of lipid dysregulation, it may also point toward a more specific mechanism. For instance, PFOA has been reported to induce oxidative stress [U.S. Environmental Protection Agency, 2025], which can, in turn, activate phospholipase A2 (PLA2). The activity of this enzyme generates lysophospholipids from membrane phospholipids [Nelson and Cox, 2005], offering a potential biological pathway for the association we observed.

Both the long-chain polyunsaturated fatty acid (n3 and n6) pathway, and the lysophospholipid observations were also significantly associated with PFAS in a previous metabolomics study in mothers and children [Prince et al., 2023].

### 4.3 Chlorinated compounds

For 3,5-dichloro-2,6-dihydroxybenzoic acid, we found no significant pathway-level enrichment, suggesting limited evidence of broad metabolic disruption in this dataset.

For 4-chlorobenzoic acid, whose presence can indicate exposure to certain industrial chemicals, pesticides, or their degradants, as well as some pharmaceuticals. We found that 4-chlorobenzoic acid levels were associated with metabolites from the phosphatidylethanolamine (PE) class. PEs are critical structural components of cell membranes [Nelson and Cox, 2005] and while their turnover can be triggered by oxidative stress to xenobiotics [Klaassen, 2018] the lack of associations with key oxidative stress markers makes this explanation less likely. Similarly, xenobiotic-induced membrane remodeling would be expected to affect multiple phospholipid classes, and shared hepatic processing loads would provide associations across multiple metabolic pathways.

Notably, both 3,5-dichloro-2,6-dihydroxybenzoic acid and 4-chlorobenzoic acid are benzoic acid derivatives. However, pathways containing metabolites typically involved in the detoxification of benzoic acid, such as fatty acid metabolism (acyl glycine) and benzoate metabolism, were not enriched in our analysis.

Finally, while research on 4-hydroxychlorothalonil itself is scarce, its parent compound, the fungicide chlorothalonil, has been studied. It has been investigated as a possible carcinogen and recognized endocrine disruptor, leading to its ban in the EU in 2019 [Mnif et al., 2011, Guillante et al., 2024, Union, 2019]. Interestingly, in our analysis 4-hydroxychlorothalonil was associated with the androgenic steroids class. We also found associations with the alpha-linolenic acid metabolism class, and various long chain fatty acid classes (monounsaturated, saturated, polyunsaturated n3 and n6). These connections are less direct from a literature standpoint but are plausible based on chlorothalonil’s suggested disruption of mitochondrial metabolism [Slaninova et al., 2009].

### 4.4 Other compounds

Exposure to 2,4-di-tert-butylphenol (2,4-DTBP) can originate from plastic food packaging and consumer products, although it is also naturally present in certain foods and the environment. Recent research suggests it may be an endocrine disruptor [Ren et al., 2023]. While we did not find evidence of associations with hormonal pathways like androgenic steroids, our analysis showed statistically significant relationships with the monoacylglycerol, diglycerol, phosphatidylcholine (PC), phosphatidylethanolamine (PE), and the long chain polyunsaturated fatty acid (n3 and n6) classes. This pattern, suggesting a broad disruption of fatty acid handling and membrane lipid composition, appears consistent with toxicological evidence suggesting 2,4-DTBP as a potential obesogen [Ren et al., 2023]. A proposed mechanism involves the activation of the retinoid X receptor (RXR), which is known to promote lipogenesis and adipogenesis [Ren et al., 2023]. Our findings could be interpreted as support for this mechanism, as the enrichment of mono- and diacylglycerols may reflect an increased synthesis of triglycerides [Nelson and Cox, 2005], while the enrichment of PC and PE could indicate a higher demand for new cell membranes during adipogenesis [Nelson and Cox, 2005].

Lastly, we studied 2-naphthol sulfate, a phase II detoxification metabolite. Higher levels of this metabolite indicates higher levels of exposure to naphthalene which is a polycyclic aromatic hydrocarbon (PAH) found in sources such as mothballs, industrial and traffic emissions, and tobacco smoke. Naphthalene is recognized as a carcinogen in animals, and has been under investigation for potential carcinogenicity in humans [IARC, 2002]. We found associations of 2-naphthol sulfate with the lysophospholipid, benzoate metabolism, plasmalogens, and xanthine metabolism classes. These associations may reflect the consequences of naphthalene-induced oxidative stress [IARC, 2002]. The links to plasmalogens (which are consumed by oxidative attack) [Voet et al., 2016] and lysophospholipids (byproducts of membrane damage)[Nelson and Cox, 2005] suggest significant lipid peroxidation. Concurrently, the xanthine metabolism association points to a potential source of this oxidative burden, as xanthine oxidase is a primary generator of reactive oxygen species [Voet et al., 2016]. Finally, the benzoate metabolism link may reflect the phase II detoxification process.

### 4.5 Strengths

This study addresses several key limitations that have constrained previous epidemiological investigations of environmental exposures and metabolic responses. First, we investigated the metabolic response of many exposures simultaneously, thereby reducing the risk of selective reporting. Second, we employed left-censored regression models to rigorously handle non-detect values and technical variation in metabolomic data, facilitating faithful representation of exposure-metabolite relationships. Third, alongside our metabolic platform’s broad coverage, the metabolomic annotations underlying our pathway analyses represent a large number of high-confidence identifications. Fourth, we applied a relatively stringent multiple-testing correction procedure, limiting the likelihood of false-positive associations. Lastly, we leveraged state-of-the-art spatiotemporally resolved air pollution exposure surfaces from the EXPANSE consortium. These methodological considerations collectively enable more comprehensive and reliable characterization of the biological pathways linking environmental exposures to human health outcomes.

### 4.6 Limitations

Several important limitations should be considered when interpreting these findings. Some of these apply to all metabolome-wide association study (MWAS) approaches in general, while others are more specific to our study design and implementation. MWAS approaches like ours employ standardized adjustment models that, while consistent across exposures, may not capture the unique confounding structures specific to each environmental factor examined. Moreover, like almost all other MWAS studies, we were limited to metabolic information at a single time point, which provides only a snapshot of what are inherently dynamic biological processes. In reality, metabolic responses to environmental exposures can be more complicated, involving temporal fluctuations in both exposure levels and biological responses, potential adaptation mechanisms over time, and cumulative effects that may not be captured by cross-sectional measurements. Similarly, despite the broad coverage by the four chromatographic separations methods from the Metabolon platform, for many metabolic pathways only a subset of the involved metabolites could be measured – potentially missing key regulatory molecules or alternative pathway branches that could provide a more complete picture of the biological responses to environmental exposures. Lastly, these results are based on circulating plasma metabolites, which may not capture internal disruptions in pathways whose metabolites are not readily exported into the exometabolome, which can result in false negative findings [Lee et al., 2025, Cooke et al., 2025].

Other limitations are more specific to our study. The considerable time period over which plasma collection took place (spanning multiple years from 2002-2019) may have increased the risk of unobserved confounding due to general temporal trends, including shifts in population behavior or environmental regulations, and evolving exposure patterns that could bias our exposure-metabolome associations. The association of some air pollutants with the food component metabolite class may indicate such residual confounding. Additionally, our analysis represents a secondary analysis of data originally collected for a study aiming to study the effect of genetic variation on metabolite levels, which could have introduced potential selection bias. Lastly, due to the high-number of associations in the background set, the threshold for enrichment was very high in the internal exposure domain. This was especially evident for the KEGG pathways which had fewer members in general and therefore almost all members of the pathway had to be nominally statistically significant for the pathway to be considered enriched.

### 4.7 Conclusion

This exposomic analysis provides evidence that long-term environmental exposures, spanning both external sources and circulating ubiquitous anthropogenic chemicals, are measurably reflected in the human metabolic phenotype. Across exposure domains, we consistently observed perturbations involving lipid metabolism, membrane remodeling, and oxidative stress–related pathways, underscoring these as recurrent biological targets, representing the metabolic signatures of hallmarks of environmental insults [Peters et al., 2021].

For internal exposures, the widespread metabolic associations of PFAS and phenolic/chlorinated compounds support prior evidence of lipid dysregulation, endocrine disruption, and mitochondrial effects. Notably, the broad lipid perturbations associated with 2,4-DTBP align with its proposed obesogenic properties, while androgenic steroid enrichment linked to 4-hydroxychlorothalonil supports concerns about chlorothalonil’s endocrine activity.

For external exposures, ozone was linked to oxidative lipid peroxidation, NO_2_ was uniquely related to tyrosine metabolism, and PM_2.5_ to energy-metabolism and acyl-carnitine disruptions. Built-environment associations likely reflect correlated behavioral and environmental contexts rather than direct biological effects of geography.

Overall, our findings illustrate the value of integrating high-resolution exposure data with untargeted metabolomics to generate mechanistically grounded hypotheses about exposome–metabolome interactions. Because these results are observational, they should be viewed as hypothesis-generating. Future studies using longitudinal designs, repeated metabolomic profiling, and diverse exposure mixtures will be critical for establishing causality and identifying associated biological pathways.

## ACKNOWLEDGEMENTS

This work was supported by the EXPANSE and EXPOSOME-NL projects. The EXPANSE project is funded by the European Union’s Horizon 2020 research and innovation programme under grant agreement No 874627. The EXPOSOME-NL project is funded through the Gravitation program of the Dutch Ministry of Education, Culture, and Science and the Netherlands Organization for Scientific Research (NWO grant number 024.004.017). J.K, L.H and T.E were supported by the Estonian Research Council grant PRG1291. J.K and L.H were supported by the Estonian Research Council grant PRG3105. The research was conducted using the Estonian Center of Genomics/Roadmap II funded by the Estonian Research Council (project number TT17). Data analysis was carried out in part in the High-Performance Computing Center of University of Tartu.

## ETHICS

The activities of the EstBB are regulated by the Human Genes Research Act, which was adopted in 2000 specifically for the operations of the EstBB. Individual level data analysis in the EstBB was carried out under ethical approval 1.1-12/1468 from the Estonian Committee on Bioethics and Human Research (Estonian Ministry of Social Affairs), using data according to release application 3-10/GI/31961 from the Estonian Biobank.

## DATA AVAILABILITY

Data used is person-level data of Estonian Biobank participants, which cannot be published by the Estonian law. All data access to the Estonian Biobank’s data must adhere to the informed consent regulations established by the Estonian Committee on Bioethics and Human Research. Access to the datasets analysed in the current study can be arranged on a reasonable request to the Estonian Biobank. To initiate a request for phenotype data, it is necessary to submit a preliminary request to the Estonian Biobank. Information about data access, including necessary steps required to access data on the University of Tartu servers can be found at https://genomics.ut.ee/en/content/estonian-biobank.

## SUMMARY LEVEL DATA AVAILABILITY

Summary level data for all associations is available from [enter repository name and DOI] upon publication.

## CODE AVAILABILITY

Analysis scripts are available from [enter repository name and DOI] upon publication.

## CONTRIBUTIONS

M.J.O: Conceptualization, data curation, formal analysis, investigation, software, validation, visualization, methodology, writing - original draft, writing – review & editing; R.V: Supervision, conceptualization, writing – review & editing, funding acquisition, resources; K.d.H: writing – review & editing; T.E: data curation, funding acquisition, resources, writing - review and editing; L.H: data curation, formal analysis; J.V: Supervision, conceptualization, writing – review & editing; J.K: Conceptualization, data curation, investigation, methodology, project administration, supervision, writing - review and editing; E.R.T: data collection, data curation, project administration, resources.

## CONSORTIA

Estonian Biobank research team members are Lili Milani, Andres Metspalu, Tõnu Esko and Mait Metspalu.

## SUPPLEMENTARY MATERIAL

### S1 SUPPLEMENTARY METHODS I: PREPROCESSING INTERNAL EXPOSURES

In the analysis with internal exposures, we could not employ our strategy from the single metabolite analysis for external exposures that handled the nondetects and technical variation. Namely, in those analyses only the outcome had nondetects and technical variation and as such we could model this directly by e.g. including main effects for batch id in the regression model. But in the internal exposure analysis, we now have – in addition to the metabolite outcome – a left-censored predictor. This dual censoring brings its challenges, also with the estimation of technical variation that is often intertwined with the nondetects. We chose for a simple, pragmatic approach where we introduce an extra preprocessing step for the predictors in analyses involving the internal exposures before fitting the same model as for the other domains. The same preprocessing was done for the EDTA variable that was included as adjustment variable.

It was important to model the technical variation and handle the nondetects for several reasons. First, in the internal exposure analysis we are regressing the metabolites on the exogenous compounds. These measurements are potentially from the same chromatographic separation method. Therefore, without adequate adjustment for technical variation these results may be confounded by analysis methods simply because metabolites from the same method can exhibit similar technical variation.

To adequately remove the technical variation, not many nondetects could be present, because these distort the technical variation estimation in this preprocessing approach where we do no model the left-censored nature of the nondetects. Therefore, we only included internal exposures with at least 90% detects in metabolites.

Technical variation was then removed per metabolite for these analyses by fitting a mixed model with a random intercept for the batch, a random slope for the injection order within a batch, and subtracting these random effect estimates from the intensity value of the metabolite. Before running this model, potential nondetects were imputed with half minimum value batch. With the high number of detects we expect this to only have minimally distorted the technical variation estimation.

In lme4 notation this can be written as:

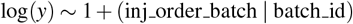

In math notation:

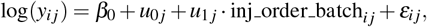

where

*i* indexes observations within batch *j*,

*j* indexes batches,

*β*_0_ is the fixed intercept,

*u*_0 *j*_ is the random intercept for batch *j*,

*u*_1 *j*_ is the random slope for injection order within batch *j*,

*ε*_*i j*_ is the residual error term.

The random effects follow a multivariate normal distribution:

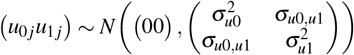

and the residual errors are

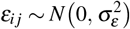

### S2 SUPPLEMENTARY INFORMATION I: PRINCIPAL COMPONENT SUMMARY

#### S2.1 Air pollution

Proportion of variance: PC1 = 62%, PC2 = 29%, PC3 = 6%, PC4 = 2%.

**Table S1.** Principal component loadings for air pollutants.

|  | PC1 | PC2 | PC3 | PC4 |
| --- | --- | --- | --- | --- |
| *NO*<sub>2</sub> | -0.50 | 0.49 | -0.55 | 0.46 |
| Ozone | 0.49 | -0.49 | -0.67 | 0.26 |
| *PM*<sub>10</sub> | -0.55 | -0.38 | -0.37 | -0.64 |
| *PM*<sub>2.5</sub> | -0.45 | -0.61 | 0.33 | 0.56 |

#### S2.2 Built environment

Proportion of variance: PC1 = 49%, PC2 = 28%, PC3 = 14%, PC4 = 6%, PC5 = 3%, PC6 = *<* 1%.

**Table S2.** Principal component loadings for the built environment. MSAVI = modified soil adjusted vegetation index; NDVI = Normalized Difference Vegetation Index; BSS = distance to nearest sea/ocean; BSI = distance to nearest inland freshwater; BSW = distance to nearest blue space; UR = Population density as focal mean within a 1500m square.

|  | PC1 | PC2 | PC3 | PC4 | PC5 | PC6 |
| --- | --- | --- | --- | --- | --- | --- |
| MSAVI | 0.53 | 0.22 | -0.26 | -0.33 | 0.01 | -0.71 |
| NDVI | 0.53 | 0.21 | -0.27 | -0.32 | -0.02 | 0.71 |
| BSI | 0.35 | -0.55 | 0.19 | 0.06 | -0.73 | -0.02 |
| BSS | 0.03 | 0.52 | 0.79 | -0.25 | -0.19 | 0.01 |
| BSW | 0.33 | -0.52 | 0.43 | -0.17 | 0.64 | 0.02 |
| UR | -0.46 | -0.25 | -0.10 | -0.84 | -0.12 | 0.00 |

### S3 SUPPLEMENTARY INFORMATION II: ASSESSMENT OF THE BLOOD METABOLOME

In total, 1505 compounds were detected. After excluding compounds with insufficient detection rates (*<* 40%), 1288 compounds remained for analysis. 688 of these compounds were detected using the C18 negative ion mode method, 292 using the C18 positive ion method optimized for hydrophilic species, 224 using the C18 positive ion method optimized for hydrophobic species, and 84 using the HILIC method for polar compounds

**Figure S1.**
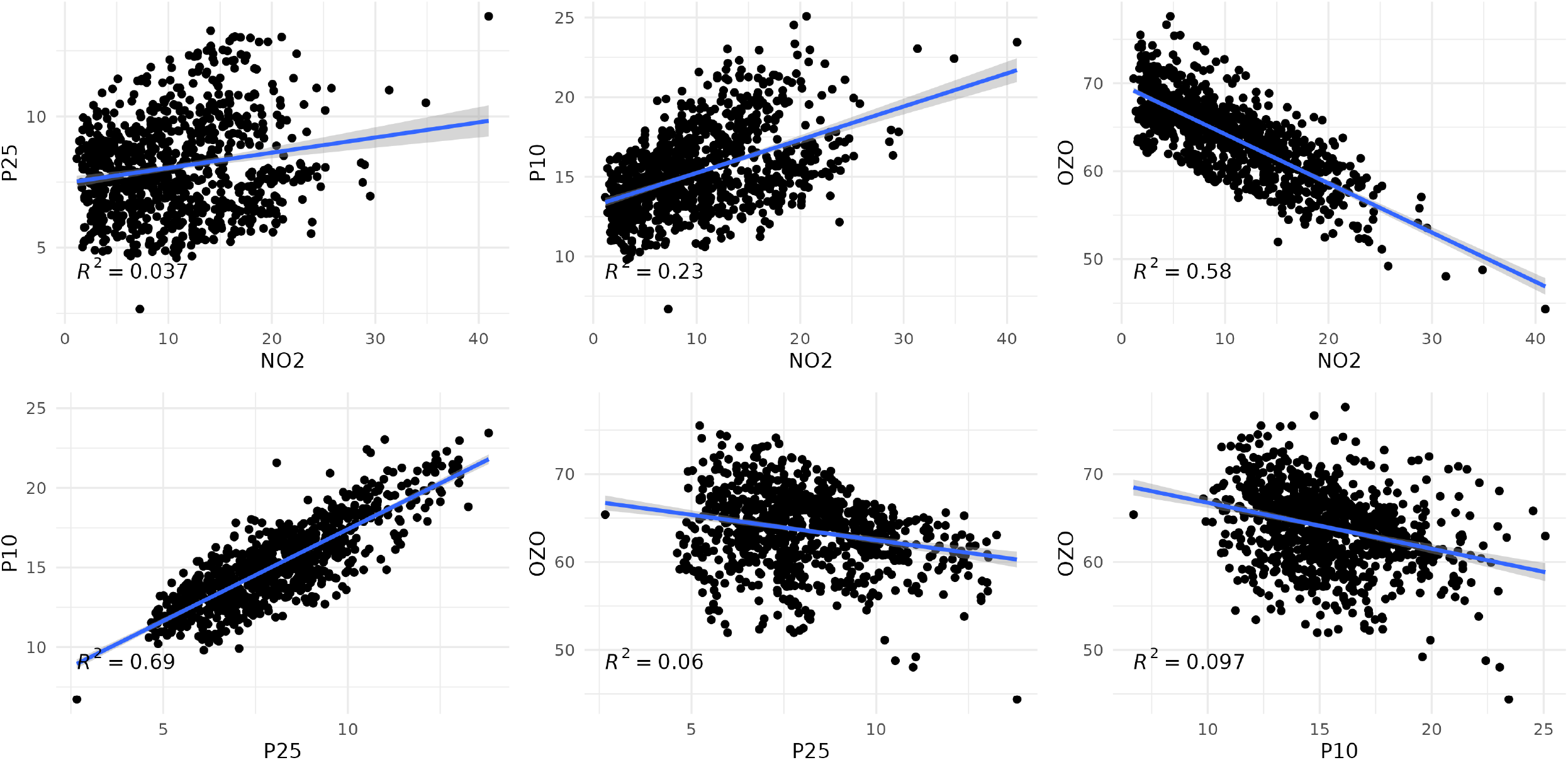
Correlation between all studied air pollutants

**Figure S2.**
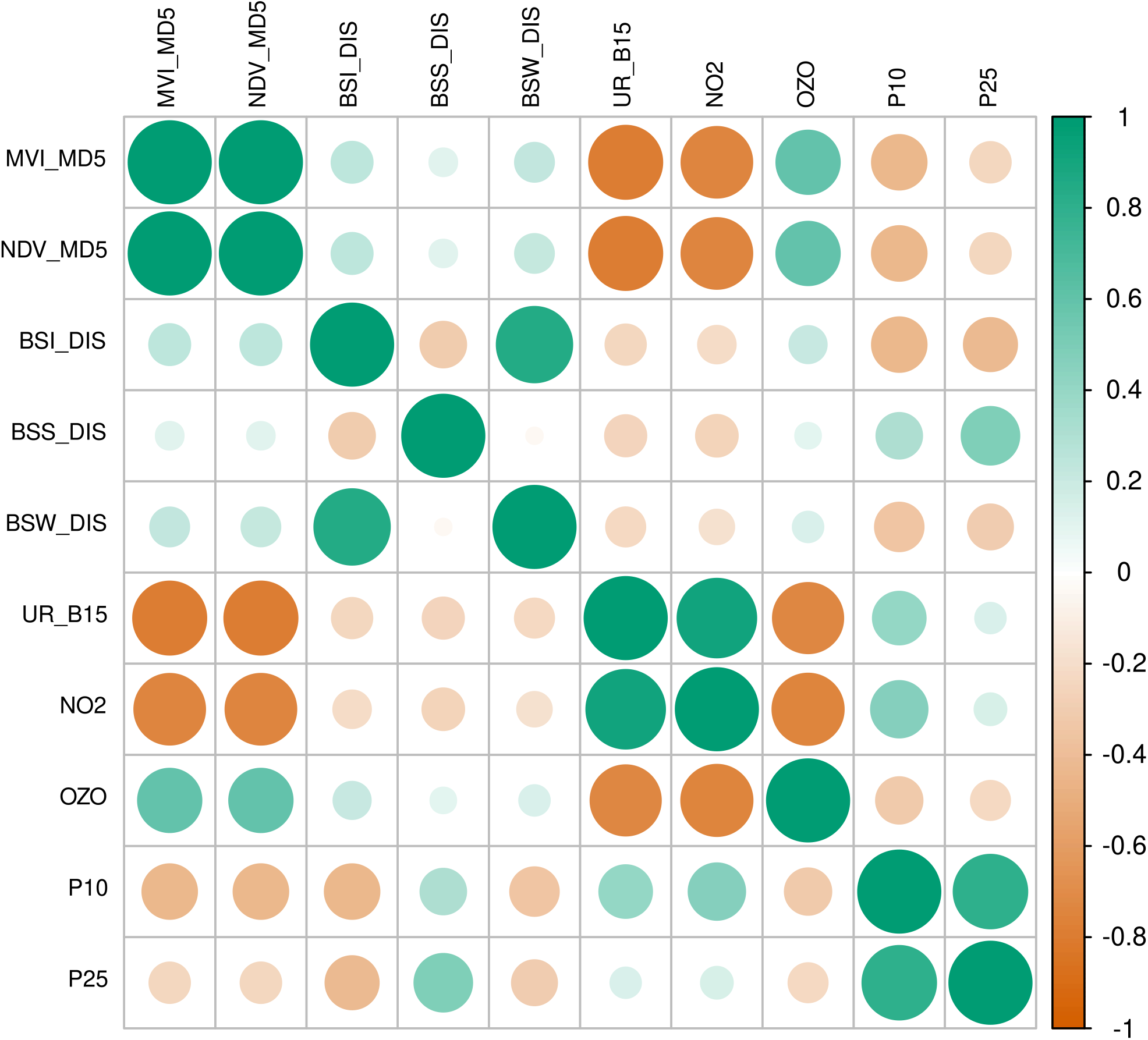
Correlation matrix (Spearman coefficient) of all external exposures. BSI = distance to nearest inland freshwater; BSS = distance to nearest sea/ocean; BSW distance to nearest blue space; MSAVI = Modified Soil Adjusted Vegetation Index; NDVI = Normalized Difference Vegetation Index

**Figure S3.**
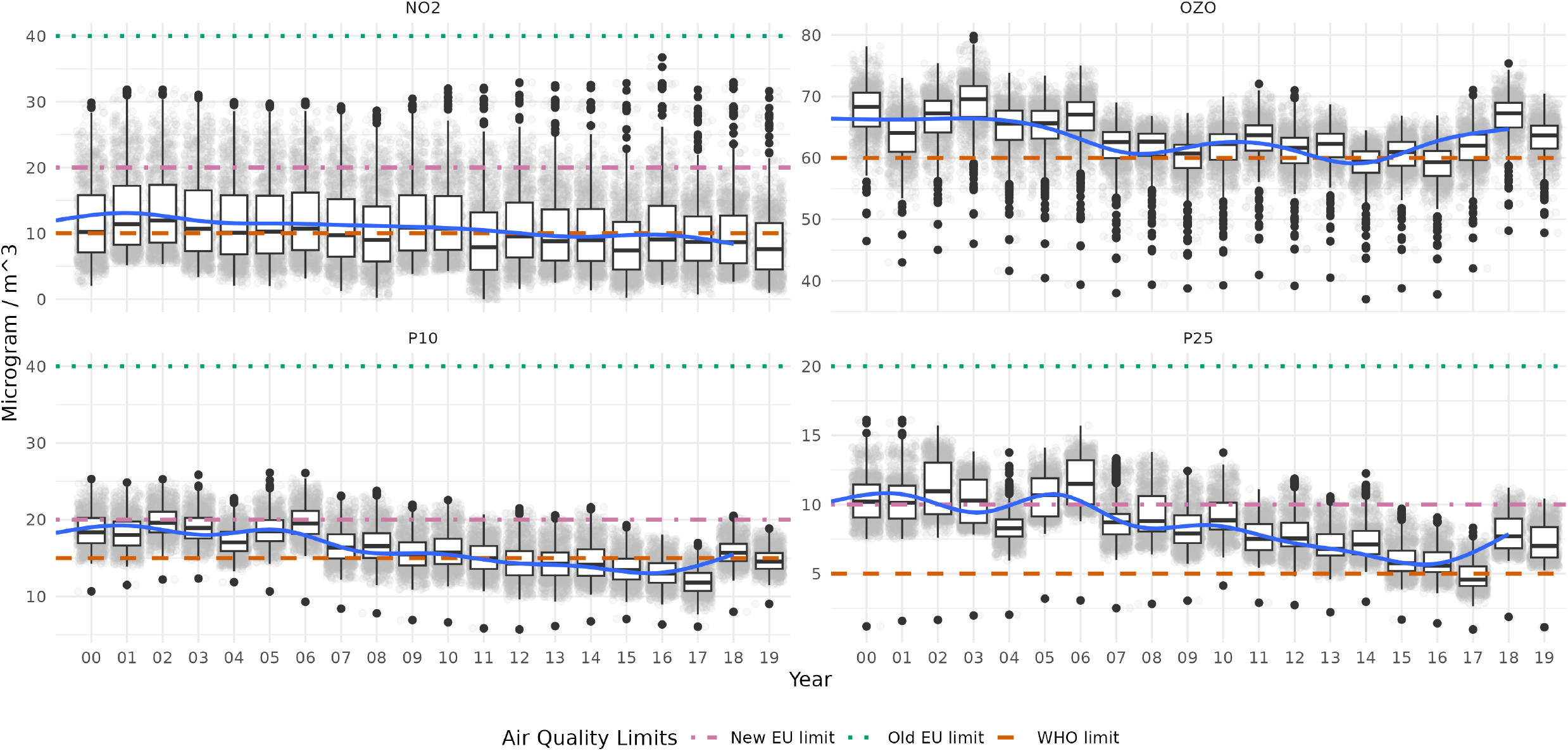
Average air pollution levels per year for the home address for participants included. The trend line is a simple regression of the air pollutant on year with cubic regression spline smooths (ggplot2 geom smooth default). The apparent increase in 2018–2019 is an artefact of more people from cities joining that year. WHO ozone limit refers to peak season limit.

**Figure S4.**
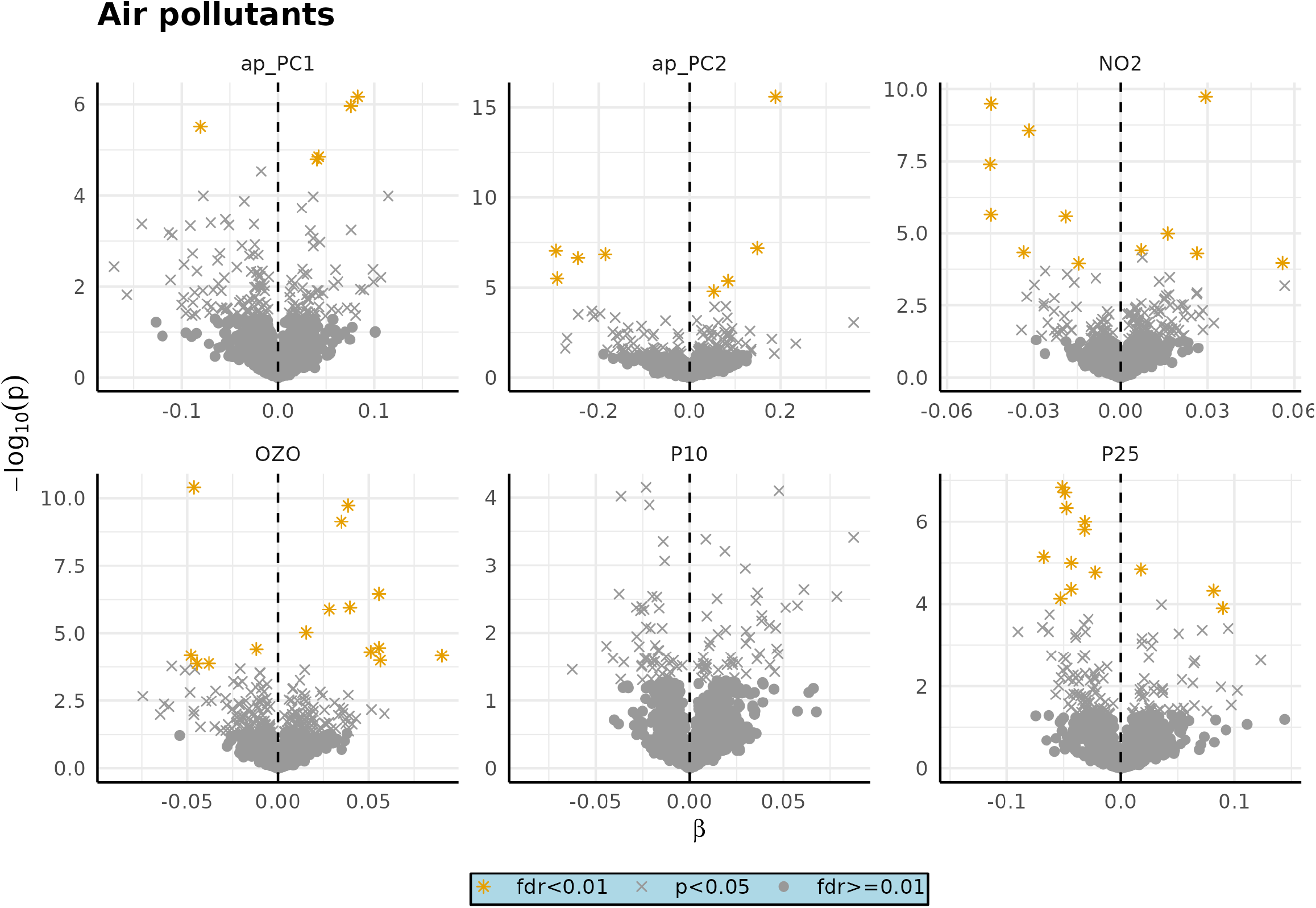
Volcano plot of air pollutant exposures in single metabolite at a time analysis.

**Figure S5.**
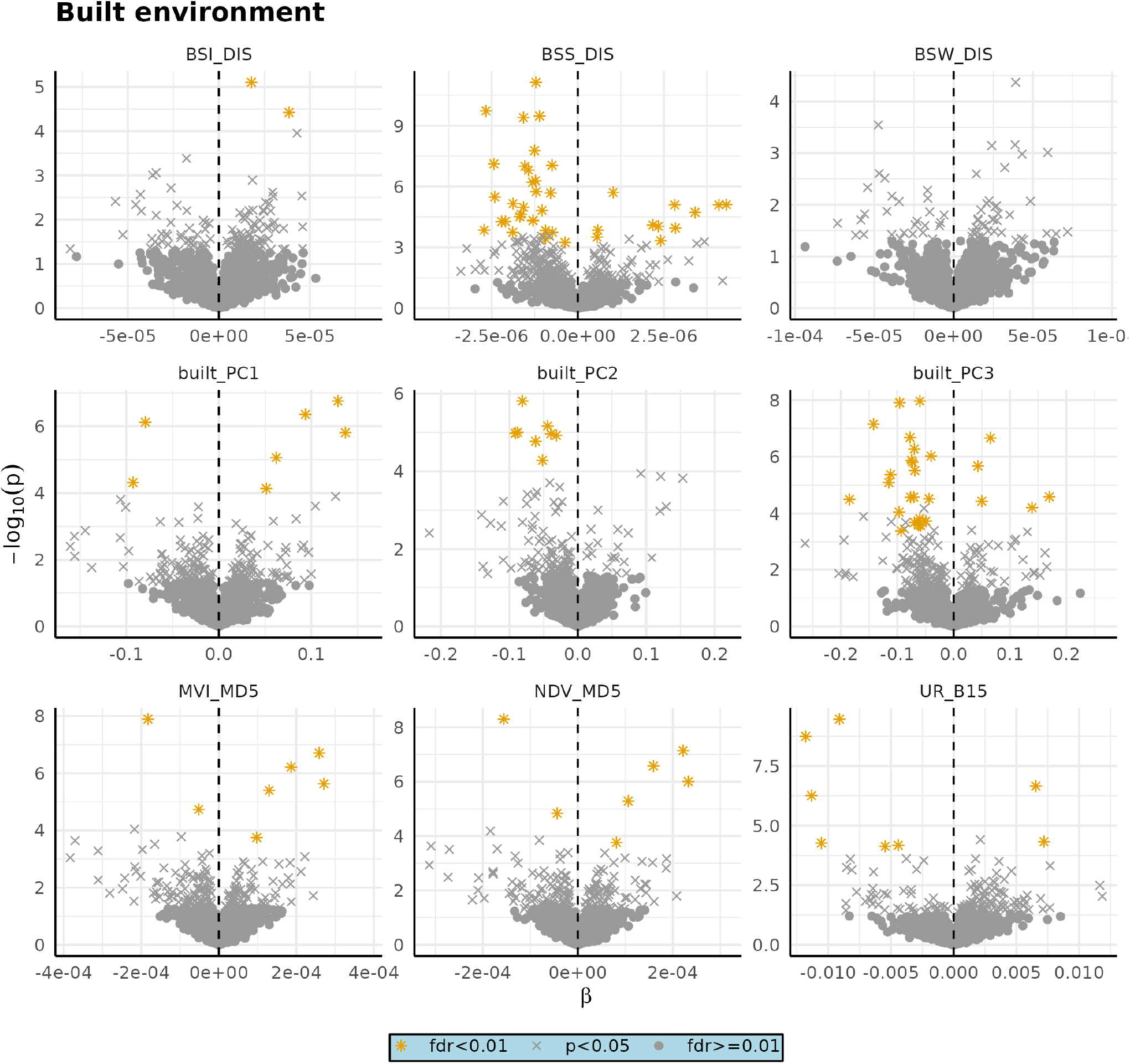
Volcano plot of built environment exposures in single metabolite at a time analysis. BSI = distance to nearest inland freshwater; BSS = distance to nearest sea/ocean; BSW distance to nearest blue space; MSAVI = Modified Soil Adjusted Vegetation Index; NDVI = Normalized Difference Vegetation Index.

**Table S3.**
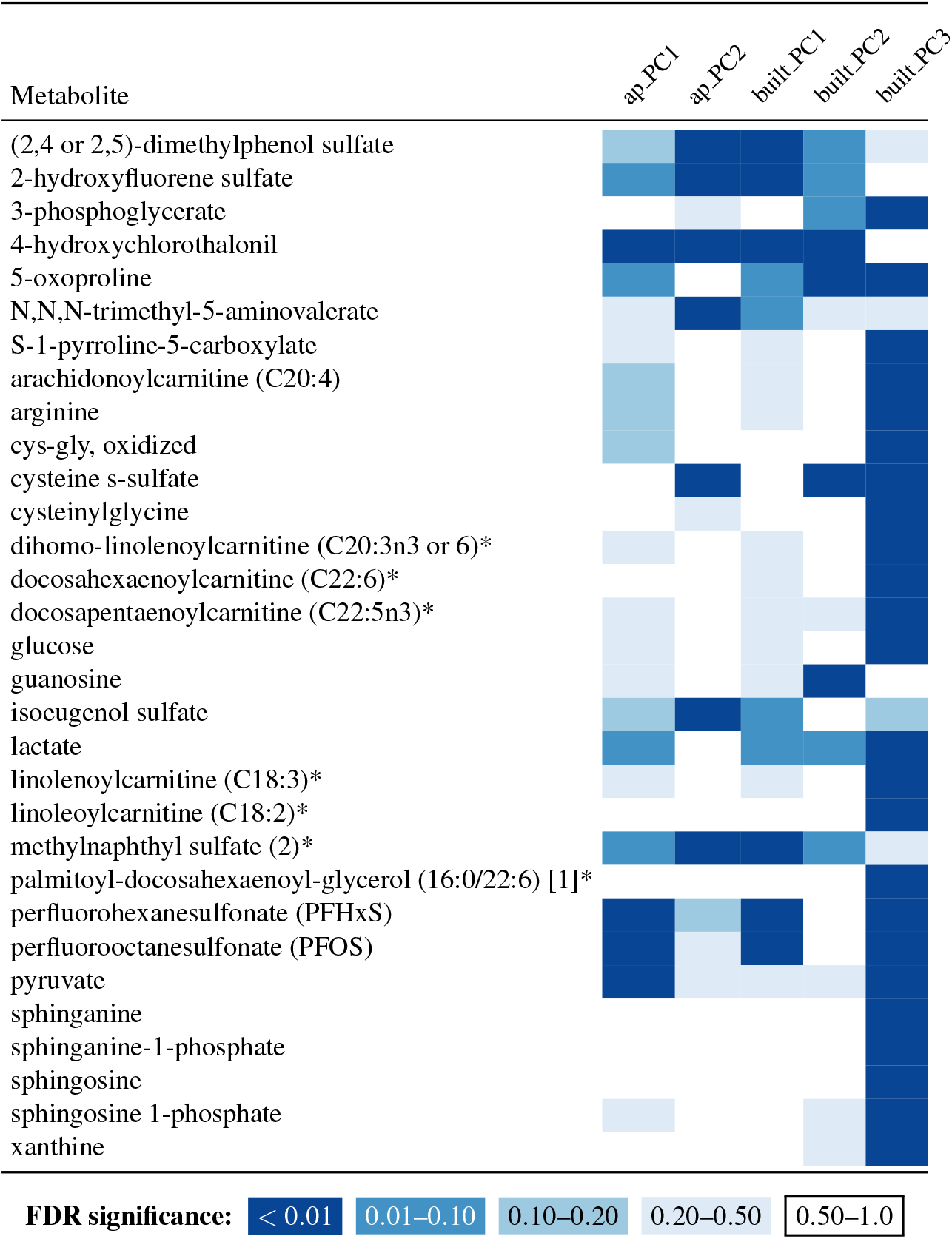
Associations of exposures with single metabolites, PCA external exposures. Metabolites that were statistically significantly associated with at least one of the presented exposures. Full biological name: compound confirmed via authentic chemical standard; high identity confidence (MSI Tier 1 identification); Biochemical Name: Compound identity remains unconfirmed by a standard, but identity confidence is high (Not Tier 1); Biochemical Name: Compound for which a chemical standard is currently unavailable, but identity or provided information is reasonably confident (Not Tier 1); Biochemical Name (#) or [#]: Indicates a structural isomer of another compound in the Metabolon spectral library (e.g., sulfated steroids or diacylglycerols with multiple possible stereospecific configurations indistinguishable by mass spectrometry). PC = principal component; FDR = false discovery rate

**Table S4.** Metabolon classes included in analysis.

| Class | Subclass |
| --- | --- |
| Amino Acid | Alanine and Aspartate Metabolism |
| Amino Acid | Glutamate Metabolism |
| Amino Acid | Glutathione Metabolism |
| Amino Acid | Glycine, Serine and Threonine Metabolism |
| Amino Acid | Histidine Metabolism |
| Amino Acid | Leucine, Isoleucine and Valine Metabolism |
| Amino Acid | Lysine Metabolism |
| Amino Acid | Methionine, Cysteine, SAM and Taurine Metabolism |
| Amino Acid | Phenylalanine Metabolism |
| Amino Acid | Polyamine Metabolism |
| Amino Acid | Tryptophan Metabolism |
| Amino Acid | Tyrosine Metabolism |
| Amino Acid | Urea cycle; Arginine and Proline Metabolism |
| Carbohydrate | Aminosugar Metabolism |
| Carbohydrate | Glycolysis, Gluconeogenesis, and Pyruvate Metabolism |
| Carbohydrate | Pentose Metabolism |
| Cofactors and Vitamins | Ascorbate and Aldarate Metabolism |
| Cofactors and Vitamins | Hemoglobin and Porphyrin Metabolism |
| Cofactors and Vitamins | Nicotinate and Nicotinamide Metabolism |
| Cofactors and Vitamins | Tocopherol Metabolism |
| Cofactors and Vitamins | Vitamin A Metabolism |
| Energy | TCA Cycle |
| Lipid | Androgenic Steroids |
| Lipid | Ceramides |
| Lipid | Corticosteroids |
| Lipid | Diacylglycerol |
| Lipid | Dihydrosphingomyelins |
| Lipid | Endocannabinoid |
| Lipid | Fatty Acid Metabolism (Acyl Carnitine, Dicarboxylate) |
| Lipid | Fatty Acid Metabolism (Acyl Carnitine, Hydroxy) |
| Lipid | Fatty Acid Metabolism (Acyl Carnitine, Long Chain Saturated) |
| Lipid | Fatty Acid Metabolism (Acyl Carnitine, Medium Chain) |
| Lipid | Fatty Acid Metabolism (Acyl Carnitine, Monounsaturated) |
| Lipid | Fatty Acid Metabolism (Acyl Carnitine, Polyunsaturated) |
| Lipid | Fatty Acid Metabolism (Acyl Choline) |
| Lipid | Fatty Acid Metabolism (Acyl Glycine) |
| Lipid | Fatty Acid, Branched |
| Lipid | Fatty Acid, Dicarboxylate |
| Lipid | Fatty Acid, Dihydroxy |
| Lipid | Fatty Acid, Monohydroxy |
| Lipid | Hexosylceramides (HCER) |
| Lipid | Long Chain Monounsaturated Fatty Acid |
| Lipid | Long Chain Polyunsaturated Fatty Acid (n3 and n6) |
| Lipid | Long Chain Saturated Fatty Acid |
| Lipid | Lysophospholipid |
| Lipid | Medium Chain Fatty Acid |
| Lipid | Monoacylglycerol |
| Lipid | Phosphatidylcholine (PC) |
| Lipid | Phosphatidylethanolamine (PE) |
| Lipid | Phosphatidylinositol (PI) |
| Lipid | Phospholipid Metabolism |
| Lipid | Plasmalogen |
| Lipid | Pregnenolone Steroids |
| Lipid | Primary Bile Acid Metabolism |
| Lipid | Progestin Steroids |
| Lipid | Secondary Bile Acid Metabolism |
| Lipid | Sphingomyelins |
| Lipid | Sterol |
| Nucleotide | Purine Metabolism, (Hypo)Xanthine/Inosine containing |
| Nucleotide | Purine Metabolism, Adenine containing |
| Nucleotide | Pyrimidine Metabolism, Cytidine containing |
| Nucleotide | Pyrimidine Metabolism, Uracil containing |
| Partially Characterized Molecules | Partially Characterized Molecules |
| Peptide | Acetylated Peptides |
| Peptide | Dipeptide |
| Peptide | Gamma-glutamyl Amino Acid |
| Xenobiotics | Benzoate Metabolism |
| Xenobiotics | Chemical |
| Xenobiotics | Food Component/Plant |
| Xenobiotics | Xanthine Metabolism |

**Figure S6.**
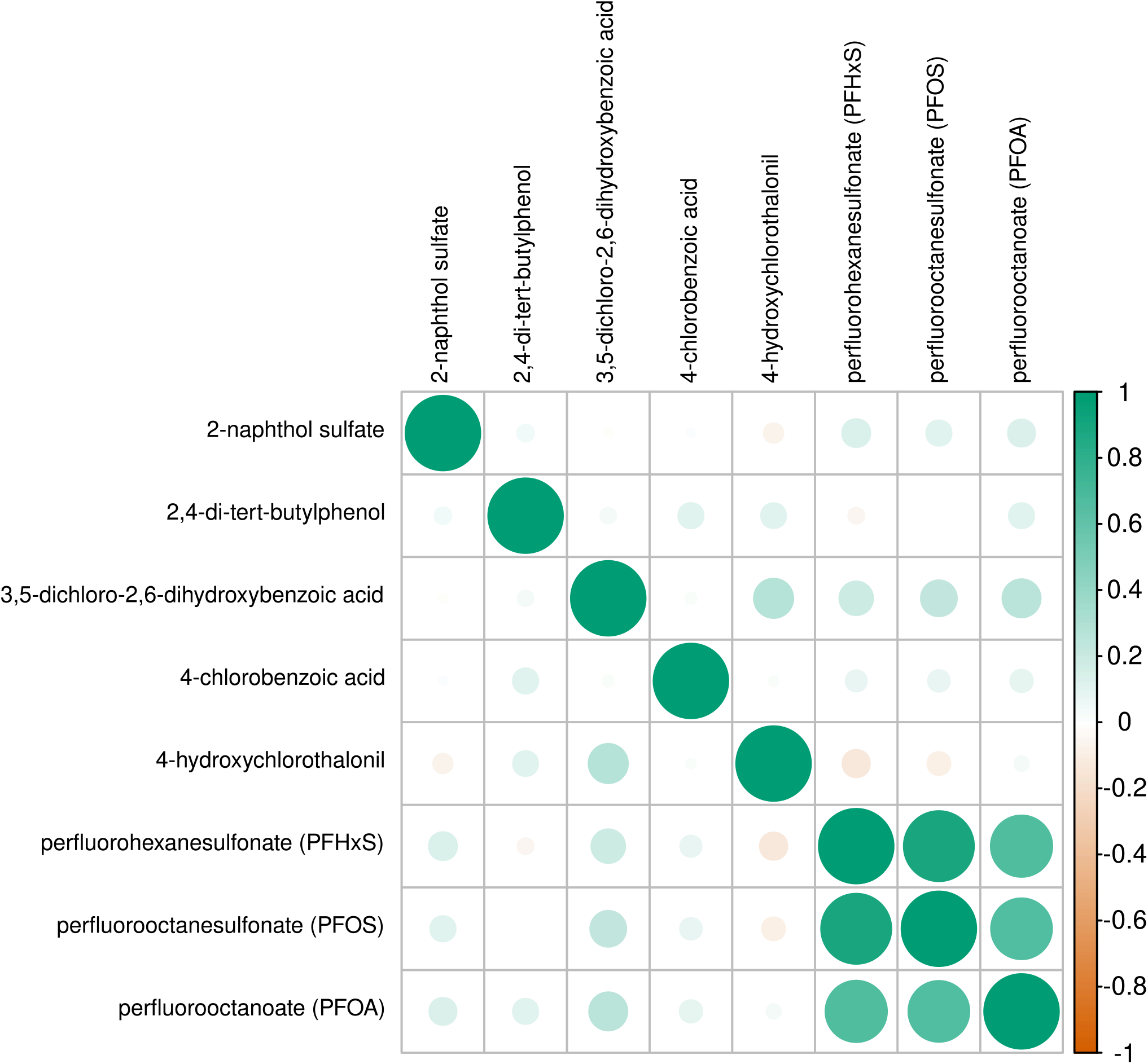
Correlation matrix (Spearman coefficient) of the ubiquitous anthropogenic chemicals.

**Figure S7.**
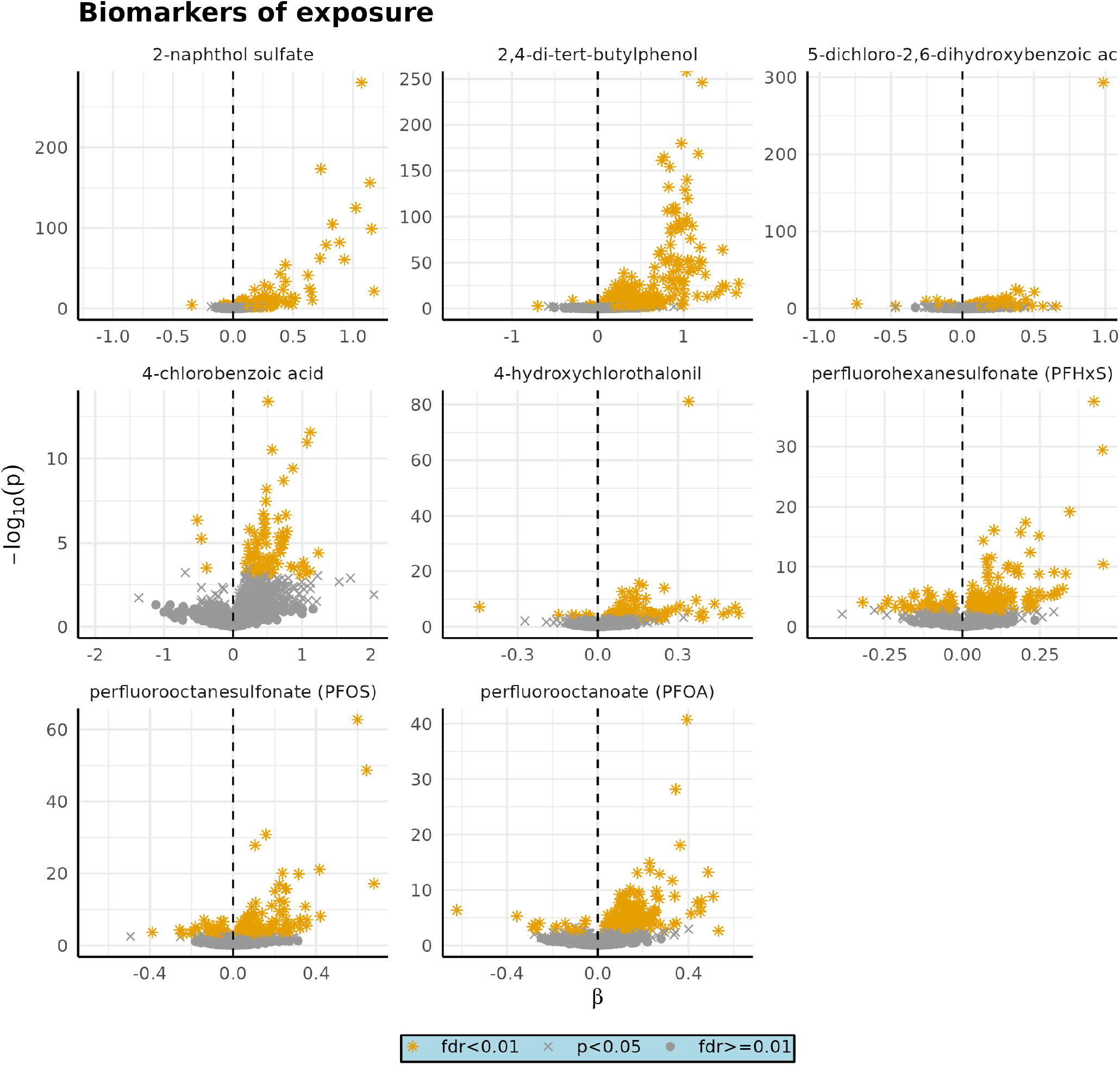
Volcano plot of internal exposures in single metabolite at a time analysis

**Table S5.**
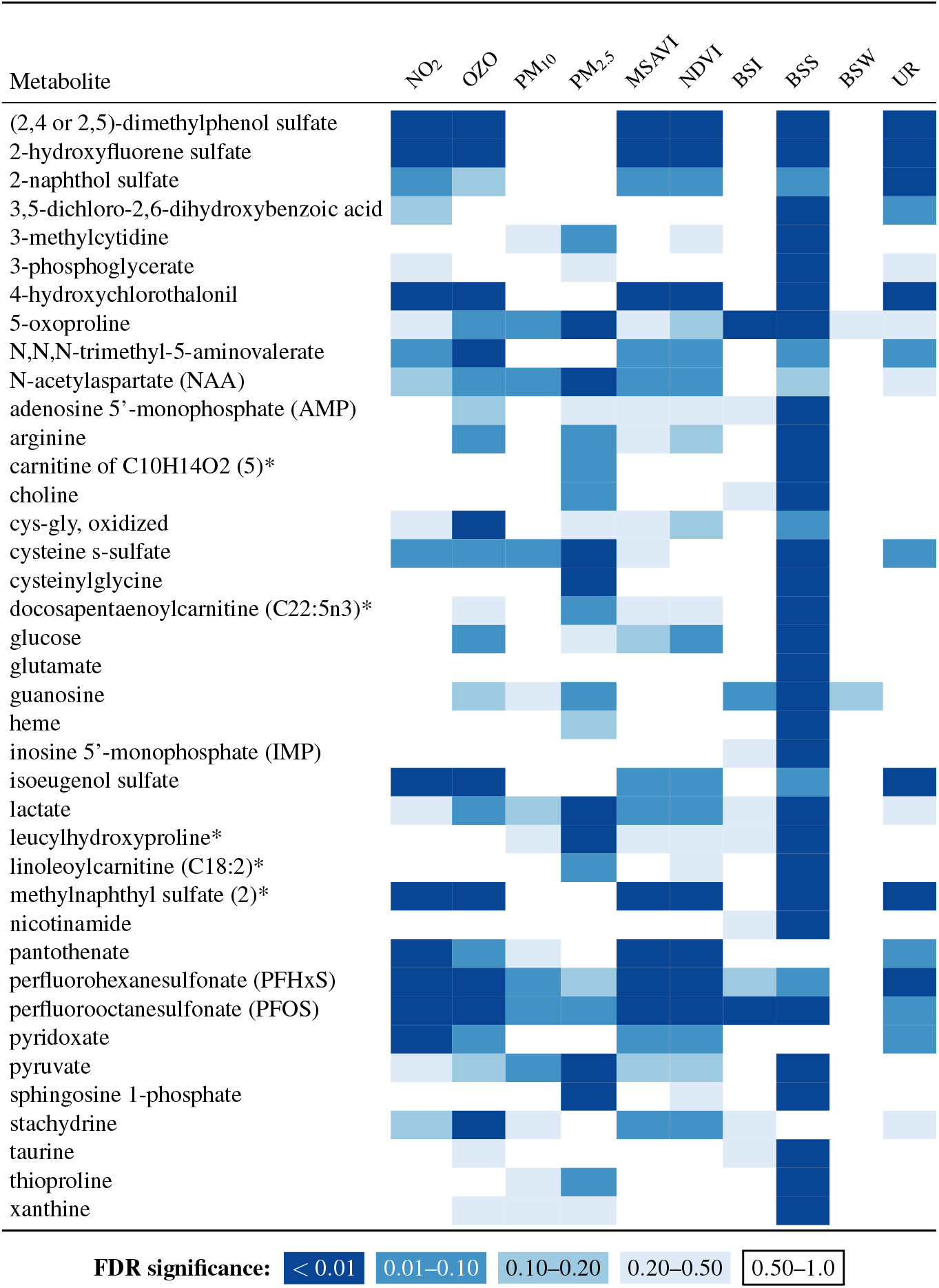
Associations of exposures with single metabolites, external exposures. Metabolites that were statistically significantly associated with at least one of the presented exposures. Full biological name: compound confirmed via authentic chemical standard; high identity confidence (MSI Tier 1 identification); Biochemical Name*: Compound identity remains unconfirmed by a standard, but identity confidence is high (Not Tier 1); Biochemical Name**: Compound for which a chemical standard is currently unavailable, but identity or provided information is reasonably confident (Not Tier 1); Biochemical Name (#) or [#]: Indicates a structural isomer of another compound in the Metabolon spectral library (e.g., sulfated steroids or diacylglycerols with multiple possible stereospecific configurations indistinguishable by mass spectrometry). FDR = false discovery rate. MSAVI = modified soil adjusted vegetation index; NDVI = Normalized Difference Vegetation Index; BSS = distance to nearest sea/ocean; BSI = distance to nearest inland freshwater; BSW = distance to nearest blue space; UR = Population density as focal mean within a 1500m square.

**Table S6.**
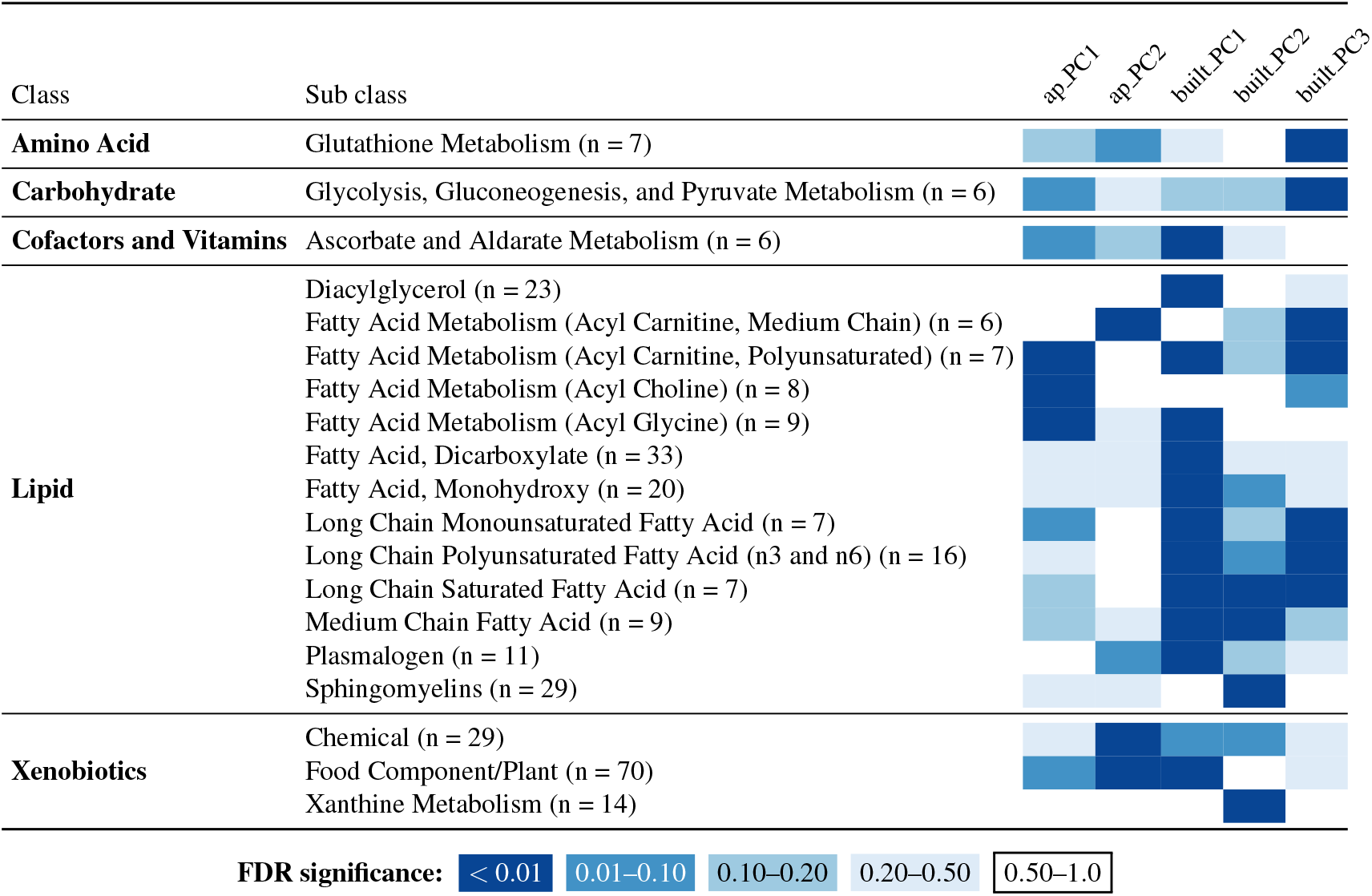
Associations of exposures with Metabolon classes, PCA external exposures. Metabolon classes that were statistically significantly related to at least one principal component of an external exposure domain. n = number of members pathway in dataset; PC = principal component; FDR = false discovery rate.

**Table S7.**
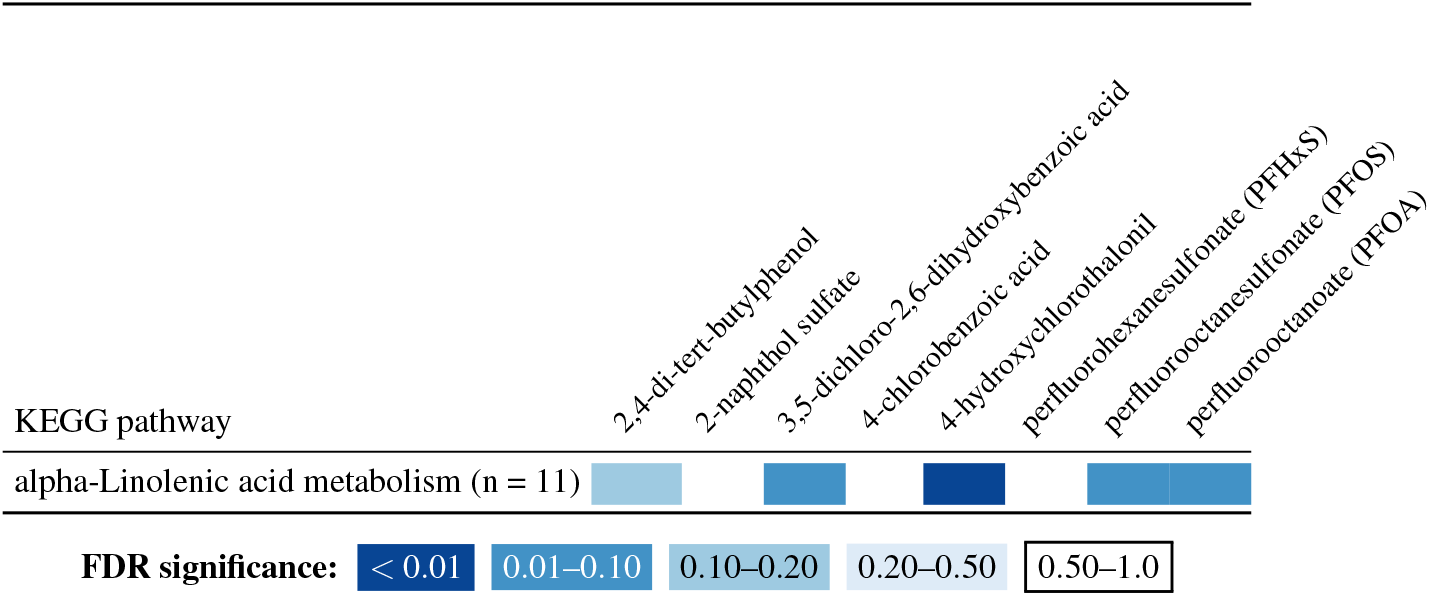
Associations of exposures with KEGG pathways, internal exposures. KEGG pathways that were statistically significantly related to at least one internal exposure. n = number of members pathway in dataset; FDR = false discovery rate; MSAVI = modified soil adjusted vegetation index; NDVI = Normalized Difference Vegetation Index; BSS = distance to nearest sea/ocean; BSI = distance to nearest inland freshwater; BSW = distance to nearest blue space; UR = Population density as focal mean within a 1500m square.

**Table S8.**
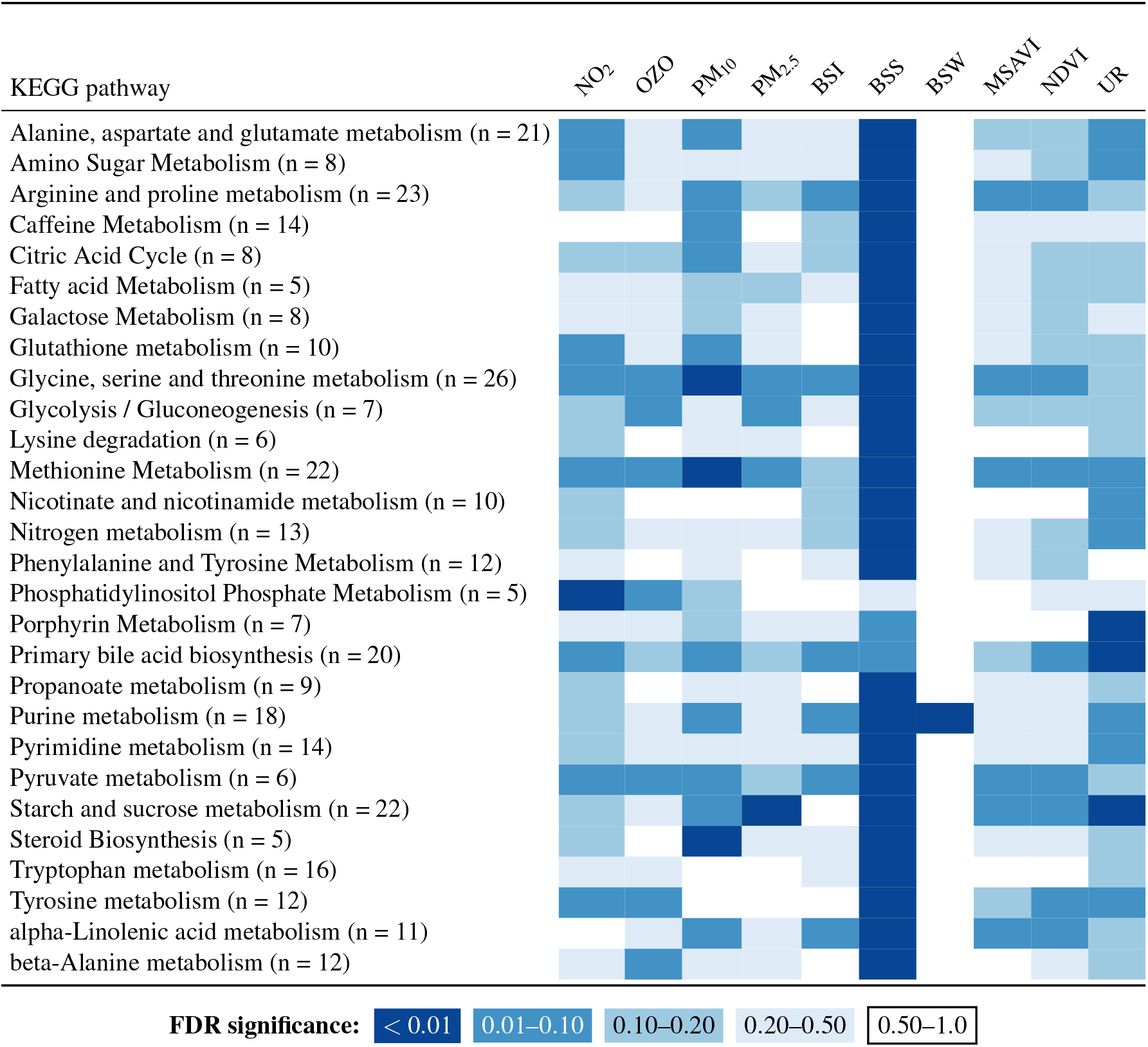
Associations of exposures with KEGG pathways, external exposures. KEGG pathways that were statistically significantly related to at least one external exposure. n = number of members pathway in dataset; FDR = false discovery rate; MSAVI = modified soil adjusted vegetation index; NDVI = Normalized Difference Vegetation Index; BSS = distance to nearest sea/ocean; BSI = distance to nearest inland freshwater; BSW = distance to nearest blue space; UR = Population density as focal mean within a 1500m square.

**Table S9.** KEGG pathways included in analysis.

| Pathway |
| --- |
| Starch and sucrose metabolism |
| Citric Acid Cycle |
| Glutathione metabolism |
| Caffeine Metabolism |
| Galactose Metabolism |
| Pyruvate metabolism |
| Nicotinate and nicotinamide metabolism |
| Methionine Metabolism |
| Histidine metabolism |
| Biosynthesis of unsaturated fatty acids |
| Amino Sugar Metabolism |
| Phospholipid Biosynthesis |
| Tryptophan metabolism |
| Phosphatidylinositol Phosphate Metabolism |
| Steroidogenesis |
| Porphyrin Metabolism |
| beta-Alanine metabolism |
| Propanoate metabolism |
| Steroid Biosynthesis |
| Fatty acid Metabolism |
| Sulfur metabolism |
| Primary bile acid biosynthesis |
| Alanine, aspartate and glutamate metabolism |
| Selenocompound metabolism |
| Purine metabolism |
| Fructose and mannose metabolism |
| Glycine, serine and threonine metabolism |
| Lysine degradation |
| Phenylalanine and Tyrosine Metabolism |
| Valine, leucine and isoleucine degradation |
| Nitrogen metabolism |
| alpha-Linolenic acid metabolism |
| Pantothenate and CoA Biosynthesis |
| Pyrimidine metabolism |
| Tyrosine metabolism |
| Arginine and proline metabolism |
| Glycerolipid Metabolism |
| Retinol Metabolism |
| Androgen and Estrogen Metabolism |
| Glycolysis / Gluconeogenesis |
| Arachidonic Acid Metabolism |

**Table S10.**
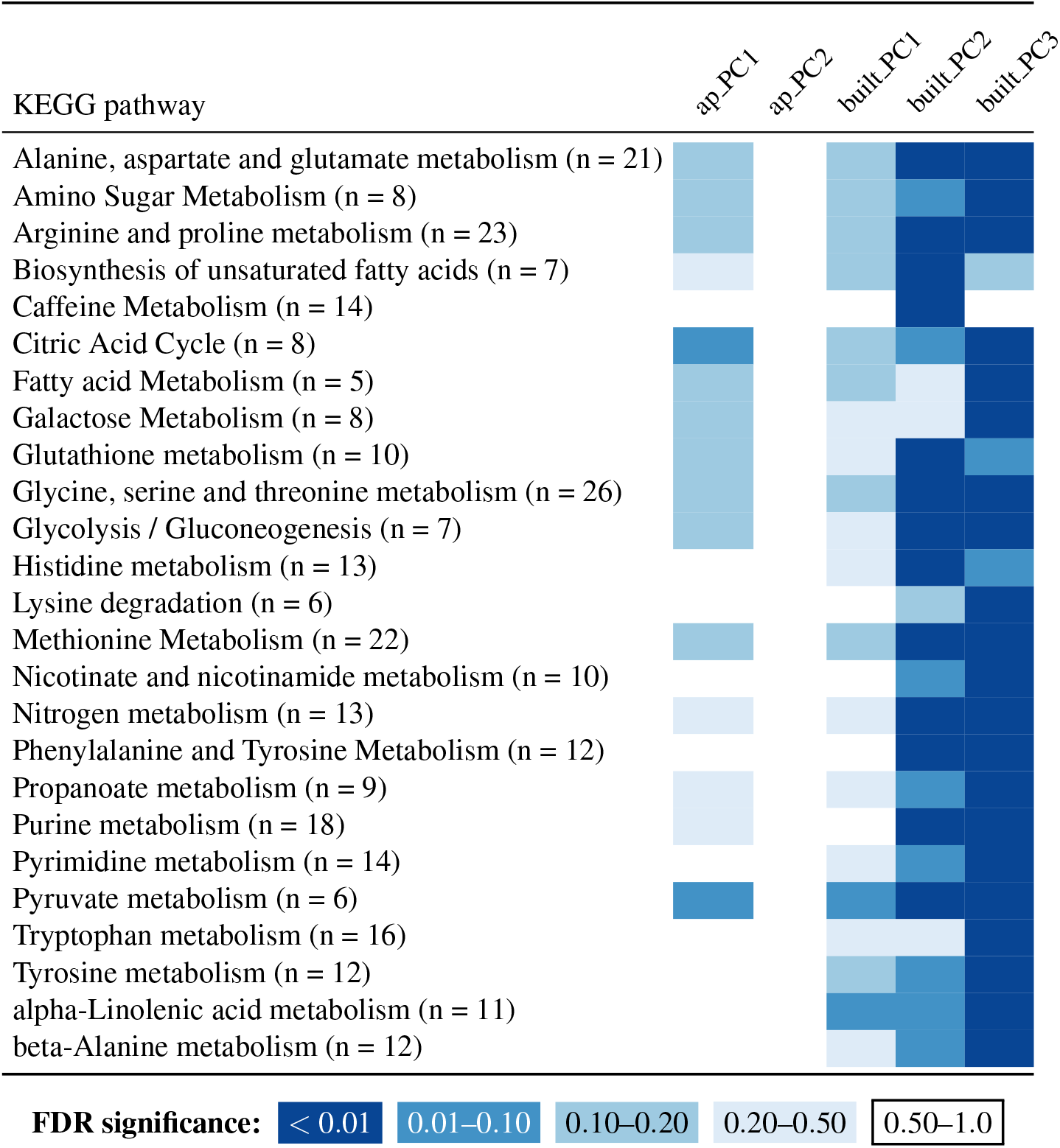
Associations of exposures with KEGG pathways, PCA external exposures. KEGG pathways that were statistically significantly related to at least one principal component of an external exposure domain. n = number of members pathway in dataset; PC = principal component; ap = air pollution; built = built environment.

